# Predicting sex, age, general cognition and mental health with machine learning on brain structural connectomes

**DOI:** 10.1101/2022.03.03.22271801

**Authors:** Hon Wah Yeung, Aleks Stolicyn, Colin R. Buchanan, Elliot M. Tucker-Drob, Mark E. Bastin, Saturnino Luz, Andrew M. McIntosh, Heather C. Whalley, Simon R. Cox, Keith Smith

**Affiliations:** Department of Psychiatry, University of Edinburgh, Edinburgh, United Kingdom; Department of Psychology, University of Edinburgh, Edinburgh, United Kingdom; Department of Psychology, University of Texas, Austin, TX, USA; Population Research Center and Center on Aging and Population Sciences, University of Texas at Austin, TX, USA; Centre for Clinical Brain Science, University of Edinburgh, Edinburgh, United Kingdom; Usher Institute, Edinburgh Medical School, The University of Edinburgh, Edinburgh, United Kingdom; Centre for Genomic and Experimental Medicine, Institute of Genetics and Molecular Medicine, University of Edinburgh, Edinburgh, UK; Department of Physics and Mathematics, Nottingham Trent University, Nottingham, United Kingdom; Lothian Birth Cohorts, University of Edinburgh, Edinburgh, United Kingdom; Scottish Imaging Network, A Platform for Scientific Excellence Collaboration (SINAPSE), Edinburgh, United Kingdom

**Keywords:** Deep Learning, Cognition, General Psychopathology, Diffusion Tensor Imaging, Structural Connectomes

## Abstract

There is increasing expectation that advanced, computationally expensive machine learning techniques, when applied to large population-wide neuroimaging datasets, will help to uncover key differences in the human brain in health and disease. We take a comprehensive approach to explore how multiple aspects of brain structural connectivity can predict sex, age, general cognitive function and general psychopathology, testing different machine learning algorithms from deep learning model (BrainNetCNN) to classical machine learning methods. We modelled *N* = 8, 183 structural connectomes from UK Biobank using six different structural network weightings obtained from diffusion MRI. Streamline count generally provided highest prediction accuracies in all prediction tasks. Deep learning did not improve on prediction accuracies from simpler linear models. Further, high correlations between gradient attribution coefficients from deep learning and model coefficients from linear models suggested the models ranked the importance of features in similar ways, which indirectly suggested the similarity in models’ strategies for making predictive decision to some extent. This highlights that model complexity is unlikely to improve detection of associations between structural connectomes and complex phenotypes with the current sample size.

## Introduction

Recent advances in neuroimaging have made it possible to acquire high quality structural brain scans in large samples, which provides opportunities for the development and application of novel machine learning (ML) techniques geared specifically for analysing brain structural architecture. Machine learning methods have the ability to uncover latent features from high-dimensional imaging data which are not apparent when using more conventional statistical methods. This is thought to be particularly promising for better understanding important biological and complex behavioural information.

The structural connectome is a comprehensive description of the connectivity between different brain regions, defined by the structure of white-matter fibre tracts (Bullmore and Sporns, 2009). The connectome is a relatively recent development which holds promise for discovery. One might expect deep learning methods to be well suited for the analysis of connnectomes, given their complexity and high-dimensionality. The structural connectome is based on information from diffusion magnetic resonance imaging (dMRI), which measures the directional diffusion of water molecules in the brain to identify connections between distal brain regions and enables estimation microstructural properties in brain white matter. The measures acquired through dMRI can be used to reconstruct individual participant connectomes in the form of adjacency matrices, where row and column indices specify to each brain region and the matrix entries are the connectivity strengths (which can be measured using various diffusion parameters) between any two regions of interest (ROI). The weighted networks constructed from different connectivity modalities then represent different interpretations of connection strength (Agosta et al., 2014; Buchanan et al., 2020; Collin et al., 2014; Hagmann et al., 2008; Jiang et al., 2020b; Robinson et al., 2010; Rutland et al., 2019).

The most commonly used connectivity weightings are based on streamline count (SC), fractional anisotropy (FA) and mean diffusivity (MD). SC provides both intra-regional as well as inter-regional streamline densities (Hagmann et al., 2008), FA measures the degree of directional dependence of the water molecular diffusion (Basser and Pierpaoli, 2011; Robinson et al., 2010), and MD measures the average magnitude of diffusion of water molecules in all directions (Agosta et al., 2014; Alexander et al., 2007; Collin et al., 2014). In addition, three newer network weightings are available thanks to the development of Neurite Orientation Dispersion and Density Imaging (NODDI) (Zhang et al., 2012). NODDI estimates include neurite density (intra-cellular volume fraction; ICVF), extra-cellular water diffusion (isotropic volume fraction; ISOVF) and tract fanning/complexity (orientation dispersion; OD).

There is accruing evidence that differences in connectomic properties between people, particularly for global network measures, are associated with basic demographic variables namely age and sex, both of which are important predictors of brain health (Buchanan et al., 2020; Madole et al., 2021; Ritchie et al., 2018). Connectomic differences are also associated with psychiatric and neurological brain disorders, and that some connectome alterations are shared across multiple brain disorders (de Lange et al., 2019; Korgaonkar et al., 2014; Ma et al., 2020). While univariate statistical methods allow assessing relationships between measures of interest and brain disorders, machine learning (ML) approaches allow combining information from different features for diagnostic classification, prediction of symptom severity levels and prediction of treatment response. More recently, researchers have been exploring ways of applying ML to connectome data. Some examples include the development of the Connectome-based Predictive Modelling approach and application of the Support Vector Machines for predicting cognitive ability, behavioral measures and clinical outcomes (Finn et al., 2015; Gong and He, 2015; Griffa et al., 2013; Jiang et al., 2020a; Payabvash et al., 2019).

Although an individual connectome provides a lower-dimensional representation of the brain when compared to a voxel-wise MRI image, there are still thousands of features - connections between brain regions - which may lead to over-fitting. To reduce dimensionality, global graph-theoretic metrics are commonly derived from connectomes and analysed as coarse representations of the participants’ brain networks (de Lange et al., 2019; Sun et al., 2017; Suo et al., 2018). A popular alternative technique is subnetwork analysis, where brain regions in the connectome are limited to those previously shown as relevant to the clinical traits or conditions of interest (Chen et al., 2016; Zheng et al., 2019). It is also common to employ feature selection to reduce dimensionality (Shen et al., 2017). Some of the common prediction models used with high-dimensional connectomic data are LASSO regression (Madole et al., 2021; Sripada et al., 2019; Wager et al., 2013), ridge regression (Gao et al., 2019; Siegel et al., 2016), elastic-net (Rahim et al., 2017), multiple kernel learning Support Vector Machine (MKL-SVM) (Xu et al., 2020), as well as relevance vector regression (Gong et al., 2014). Most of these models only consider linear interactions between ROIs and treat each connection independently. More recent studies, however, have shown that brain regions interact nonlinearly, and thus these approaches may not be optimal (Breakspear, 2017; Ocker et al., 2017; Wang et al., 2019).

Measures of cognitive ability and psychiatric symptoms, like imaging data, can be high dimensional and have often been analysed as general factors to reduce dimensionality (Cronbach and Meehl, 1955). A general intelligence factor (*g*-factor) can be derived from a sufficiently broad domain of cognitive tasks, and *g*-factors from different sets of cognitive tasks have been shown to be highly correlated (Johnson et al., 2004, 2008). Likewise, researchers have explored the possibility of a single dimension (*p*-factor) to measure one’s mental health (Caspi et al., 2014; Caspi and Moffitt, 2018; Lahey et al., 2012). Empirical evidence indicates a continuum of symptoms across multiple mental illnesses and an overlap of symptoms across mental disorders (Kotov et al., 2017), with some disorders sharing the same set of risk factors and biomarkers (Goodkind et al., 2015; Pinto et al., 2017). While molecular levels of analysis support the *g*-factor as a unitary construct, less molecular evidence provides support for the unitary nature of the *p*-factor (de la Fuente et al., 2021; Grotzinger et al., 2021, 2020). Rather, the *p*-factor may represent an emergent phenomenon, arising from the aggregation of varied mechanisms present across subsets of psychiatric conditions. Studies have found that both the *g*-factor and the *p*-factor are highly heritable in adulthood (Allegrini et al., 2020; Grotzinger et al., 2019; Harden et al., 2020), and associated with variation in the human brain connectome (Lund et al., 2020; Madole et al., 2021). The measure of general intelligence and psychopathology has been found to be strongly associated with long term outcomes (Calvin et al., 2011; Caspi et al., 2020; Deary, 2008; Plana-Ripoll et al., 2019; Strenze, 2007). The discovery of ML models that can accurately predict cognitive functioning and mental health will be beneficial to advances in the application of ML to predict different types of neurological disorders.

Rapid growth in availability of parallel computing in the recent years has enabled application of deep learning (DL). DL is very effective in extracting latent features and non-linear patterns from complex data (Vieira et al., 2017). A 3D convolutional neural network (CNN) approach has been used to investigate automated brain disease classification based on or voxel-based morphometric (VBM) features derived from structural MRI scans (Cole et al., 2017; Hu et al., 2020; Zou et al., 2017). Additionally, different graph-based neural network (GNN) models have been developed for predictive modelling based on connectivity matrices from diffusion MRI (dMRI) and resting state functional MRI (rs-fMRI) (Kawahara et al., 2017; Li and Duncan, 2020). In a previous study (Yeung et al., 2020), we have shown that the BrainNetCNN neural network architecture proposed by Kawahara et al. (2017) was more appropriate for sex classification based on brain connectome adjacency matrices compared with a naive image-based CNN architecture. With the customised CNN layers, the BrainNetCNN could potentially capture non-linearities in connectomic data that could be related to complex phenotypes.

Although there are several theoretical merits in the application of DL methods for understanding the neurobiological correlates of important between-person differences, the use of promising cutting-edge DL methods and its quantitative benefits beyond more conventional statistical methods remain moot. He et al. (2020) reported that Kernel Ridge Regression achieved comparable results with DL models (He et al., 2020). Schulz et al. (2020) also showed that non-linear models did not outperform simple linear models in predicting common phenotypes from brain scans (Schulz et al., 2020). In contrast, Abrol et al. (2021) found in their study that DL models substantially outperformed standard ML methods (Abrol et al., 2021). Large sample sizes and appropriate image processing methods constitute a highly valuable test-bed in which to directly address the challenge of assessing the performance of DL methods in comparison to other methods.

In this study, we directly compare the BrainNetCNN model against four classical ML methods when applied to structural connectomes in the UK Biobank (UKB), one of the largest structural connectome samples to date, based on 6 different structural connectivity modalities. Namely, the investigated modalities were the three common network weights – SC, FA and MD - discussed in many previous studies, and three newer and less-well studied network weights – OD, ICVF and ISOVF – derived with NODDI. Our main aims were i) to compare the performances and feature robustness on prediction of sex and age as benchmark tasks, as well as general cognition (*g*-factor) and mental health (MHQ-factor) among the 6 different connectivity modalities with the DL models, ii) to test the effect of adding external predictors to DL model, iii) to investigate how the DL model used the brain features for prediction, through the use of Gradient Attribution Map, iv) to compare the DL model’s performance, feature robustness and feature importance ranking with those of classical ML methods, using prediction of sex and age as benchmark and then extending to prediction of more complex phenotypes (cognition and psychopathology).

## Results

### Prediction performances for different tasks

We first applied the BrainNetCNN classification models to predict sex. Table 1a shows the prediction accuracies and Figure 1 shows the Receiver Operator Characteristic (ROC) curve for sex for the different structural connectivity weightings. We found that SC attained the best performance (Accuracy = 86.91%) of all modalities, on average 4.1% clear of the next best, OD. We then applied the BrainNetCNN regression models to predict age. Table 1b shows the mean absolute error (MAE) and correlation between the raw predicted age and the true values for different connectivity weightings. Here the best performance was again obtained with SC (MAE = 4.245 years).

**Table 1:** Prediction performances with BrainNetCNN of four different prediction tasks based on different connectivity weightings. MD = mean diffusivity; FA = fraction anisotropy; SC = streamline count; OD = orientation dispersion; ISOVF = isotropic volume fraction; ICVF = intracellular volume fraction. Use of connectomes based on streamline counts generally led to the best predictive performance results.

| Weights | MD | FA | SC | OD | ISOVF | ICVF |
| --- | --- | --- | --- | --- | --- | --- |
| **Validation** | 78.07 (0.54) | 80.76 (0.70) | 89.18 (0.60) | 83.53 (0.51) | 82.07 (0.75) | 80.45 (0.89) |
| **Training** | 87.17 (2.35) | 89.44 (2.40) | 94.10 (1.28) | 91.60 (1.76) | 91.49 (2.70) | 91.58 (3.27) |
| **Test** | 78.15 (0.86) | 79.74 (0.82) | 86.91 (0.72) | 82.88 (0.95) | 81.82 (0.22) | 78.34 (1.03) |

| Weights | MD | FA | SC | OD | ISOVF | ICVF |
| --- | --- | --- | --- | --- | --- | --- |
| **Validation** | 4.696 (0.640) | 4.636 (0.645) | 4.153 (0.727) | 4.374 (0.691) | 4.256 (0.703) | 4.731 (0.625) |
| **Training** | 3.797 (0.785) | 3.669 (0.796) | 3.634 (0.795) | 3.550 (0.804) | 3.538 (0.806) | 3.941 (0.765) |
| **Test** | 4.640 (0.644) | 4.674 (0.636) | 4.245 (0.706) | 4.571 (0.655) | 4.363 (0.678) | 4.692 (0.626) |

| Weights | MD | FA | SC | OD | ISOVF | ICVF |
| --- | --- | --- | --- | --- | --- | --- |
| **Validation** | 0.151 (0.770) | 0.168 (0.770) | 0.227 (0.764) | 0.185 (0.768) | 0.168 (0.780) | 0.169 (0.769) |
| **Training** | 0.232 (0.764) | 0.275 (0.761) | 0.314 (0.745) | 0.315 (0.753) | 0.286 (0.758) | 0.264 (0.763) |
| **Test** | 0.138 (0.782) | 0.168 (0.780) | 0.201 (0.780) | 0.132 (0.787) | 0.155 (0.786) | 0.160 (0.782) |

| Weights | MD | FA | SC | OD | ISOVF | ICVF |
| --- | --- | --- | --- | --- | --- | --- |
| **Validation** | 0.100 (0.803) | 0.089 (0.780) | 0.117 (0.785) | 0.118 (0.794) | 0.095 (0.795) | 0.073 (0.794) |
| **Training** | 0.238 (0.797) | 0.252 (0.768) | 0.323 (0.751) | 0.218 (0.785) | 0.403 (0.785) | 0.199 (0.787) |
| **Test** | 0.116 (0.817) | 0.112 (0.792) | 0.143 (0.790) | 0.130 (0.804) | 0.112 (0.804) | 0.103 (0.805) |

(d) MHQ-factor prediction performance (correlations with mean absolute error in brackets) with BrainNetCNN for different connectivity weightings

**Figure 1:**
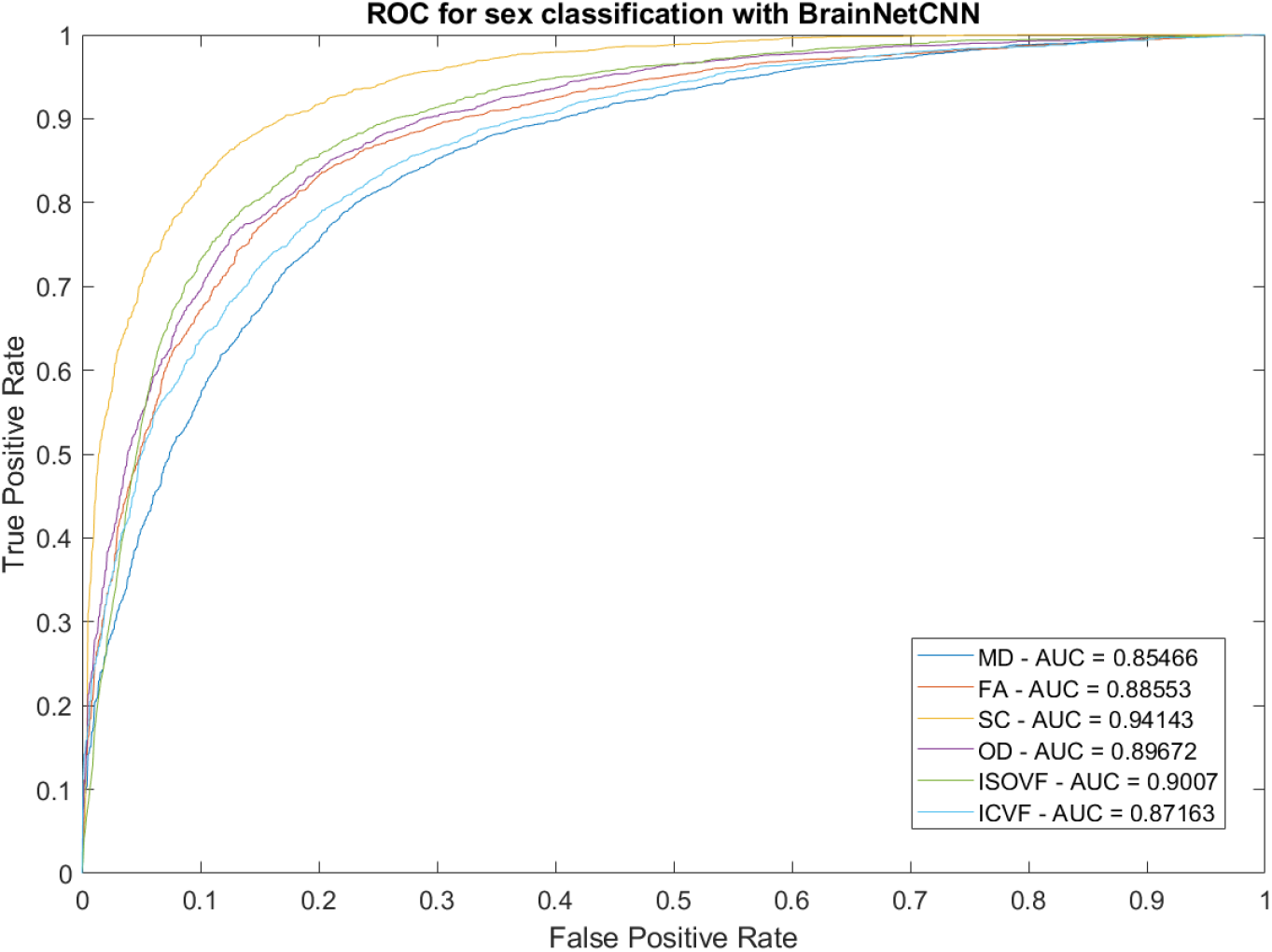
Area Under the curve (AUC) and ROC curve for sex classification with BrainNetCNN model. MD = mean diffusivity; FA = fraction anisotropy; SC = streamline count; OD = orientation dispersion; ISOVF = isotropic volume fraction; ICVF = intracellular volume fraction.

We also assessed prediction of the *g*-factor and the MHQ-factor. Table 1c shows the results for *g*-factor. Cognition and psychopathology being complex phenotypes are more subtly reflected in brain phenotypes compared to age, and hence we expected much larger prediction errors for these variables. This was indeed the case with correlation between the raw predicted *g*-factor and the true values being 60 – 75% lower compared to those observed for age prediction. Again, SC was the modality with best performance (*g*-factor: MAE = 0.780, Correlation = 0.201, MHQ-factor: MAE = 0.790, Correlation = 0.143). Table 1d shows the results for MHQ-factor, where the correlations were 80 – 85% lower compared to those observed for age prediction.

We then investigated whether addition of age and sex covariates to the DL model improved predictive performance. Age and sex were both significantly associated with the *g*-factor and MHQ-factor, and we considered that if these covariates were combined with the brain connectivity matrix, this would improve predictive performance of the DL model. Indeed, the DL model with additional covariates was able to achieve better performance for *g*-factor where all modalities achieved correlations around 0.238 - 0.251 (Table 2a). The additional covariates also improved prediction of the MHQ-factor, with all modalities achieving correlations around 0.202 - 0.245 (please see Table 2b). Moreover, when trained with additional covariates, models based on all modalities achieved comparable performances on predicting *g*-factor and MHQ-factor.

**Table 2:**
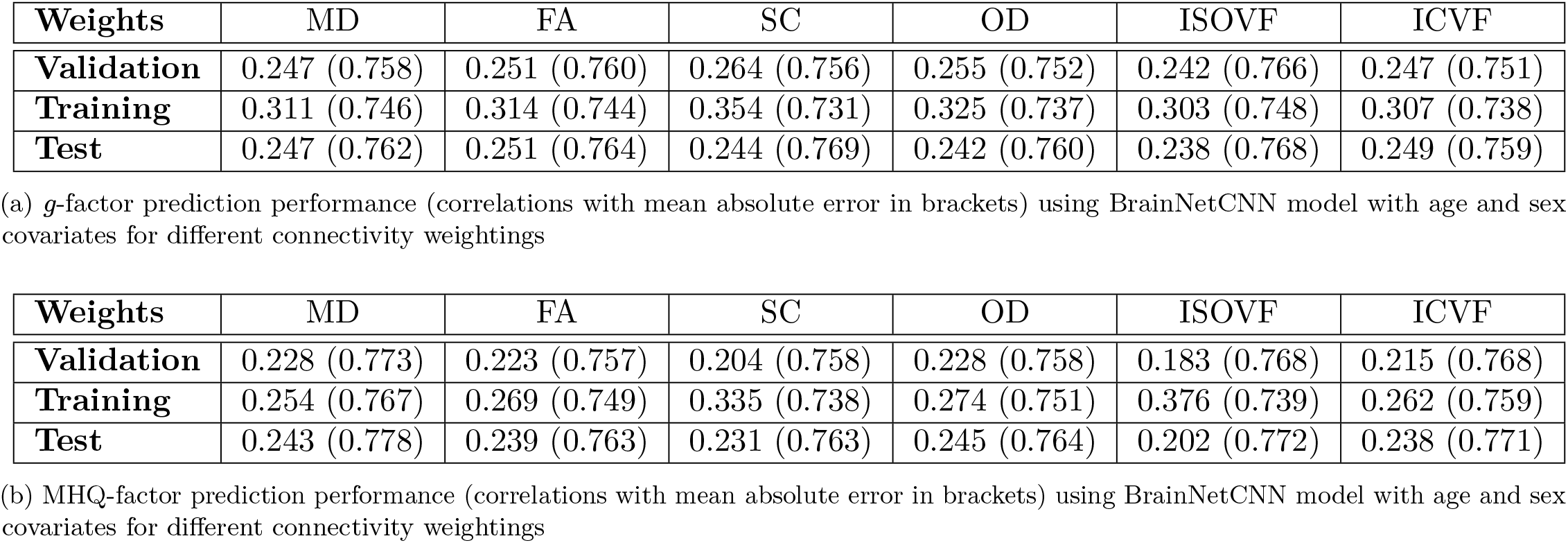
*g*-factor and MHQ-factor prediction performance using BrainNetCNN model with age and sex covariates for different connectivity weightings. MD = mean diffusivity; FA = fraction anisotropy; SC = streamline count; OD = orientation dispersion; ISOVF = Isotropic volume fraction; ICVF = Intracellular volume fraction. After adding covariates as external regressors to the BrainNetCNN model, all network weightings had comparable performances on both *g*-factor and MHQ-factor prediction.

| **Weights** | MD | FA | SC | OD | ISOVF | ICVF |
| --- | --- | --- | --- | --- | --- | --- |
| **Validation** | 0.247 (0.758) | 0.251 (0.760) | 0.264 (0.756) | 0.255 (0.752) | 0.242 (0.766) | 0.247 (0.751) |
| **Training** | 0.311 (0.746) | 0.314 (0.744) | 0.354 (0.731) | 0.325 (0.737) | 0.303 (0.748) | 0.307 (0.738) |
| **Test** | 0.247 (0.762) | 0.251 (0.764) | 0.244 (0.769) | 0.242 (0.760) | 0.238 (0.768) | 0.249 (0.759) |

| **Weights** | MD | FA | SC | OD | ISOVF | ICVF |
| --- | --- | --- | --- | --- | --- | --- |
| **Validation** | 0.228 (0.773) | 0.223 (0.757) | 0.204 (0.758) | 0.228 (0.758) | 0.183 (0.768) | 0.215 (0.768) |
| **Training** | 0.254 (0.767) | 0.269 (0.749) | 0.335 (0.738) | 0.274 (0.751) | 0.376 (0.739) | 0.262 (0.759) |
| **Test** | 0.243 (0.778) | 0.239 (0.763) | 0.231 (0.763) | 0.245 (0.764) | 0.202 (0.772) | 0.238 (0.771) |

(b) MHQ-factor prediction performance (correlations with mean absolute error in brackets) using BrainNetCNN model with age and sex covariates for different connectivity weightings

The training progress plots for the BrainNetCNN models are presented in section B.4.4 of the Supplementary Materials.

### Mapping predictive streamline count and fractional anisotropy features

Since SC and FA are the most common weights used in previous studies, the remainder of the analysis focuses on the results obtained from prediction models based on the SC and FA inputs. We computed the gradient attribution maps, averaged across folds, for the DL models to investigate how the deep neural networks made their predictive decisions, and to estimate ROI predictive powers, see Methods. Figure 2 and 3 show the circos plots of gradient maps for all the prediction tasks based respectively on the SC and FA connectomes.

**Figure 2:**
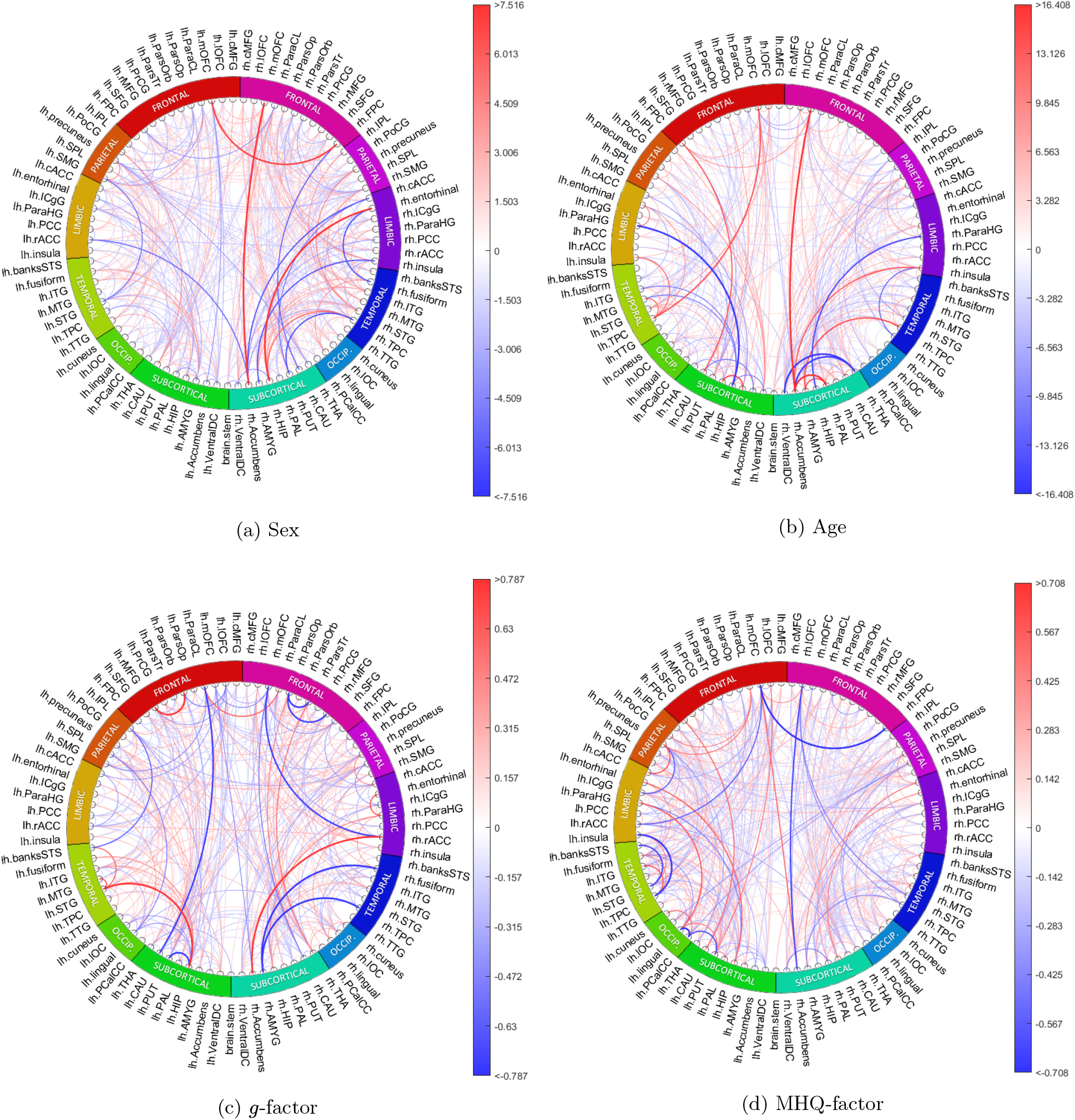
The saliency maps based on SC connectomes for all prediction tasks. SC = streamline count. Blue represents negative gradients and red represents positive gradients. The colour bar has interval of 0.1 to 99.9 percentile of the gradients

**Figure 3:**
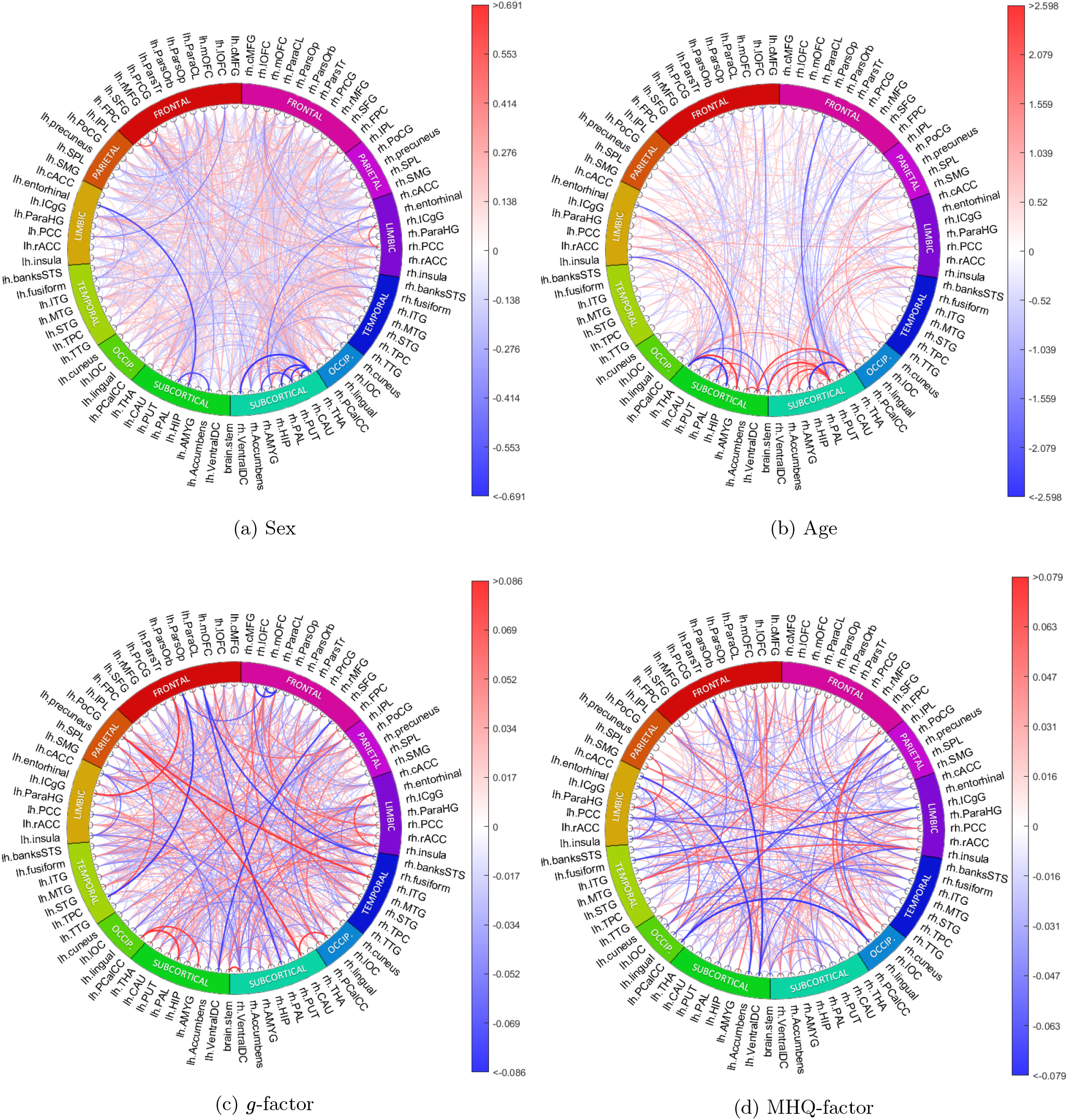
The saliency maps based on FA connectomes for all prediction tasks. FA = fractional anisotropy. Blue represents negative gradients and red represents positive gradients. The colour bar has interval of 0.1 to 99.9 percentile of the gradients

#### Sex classification gradient maps

According to the SC gradient attribution map for sex classification (SexGradient), many connections from right medial orbital-frontal, right inferior temporal regions and right hippocampus are predictive of being female (Figure 2a). Conversely, we saw that many connections from left superior temporal, right accumbens areas and left ventral diencephalon regions were chosen as being predictive of being male. In general, the precuneus, right superior temporal region, right insula, right putamen and right thalamus were considered by the model as being hubs of important connections for sex classification.

According to SexGradient based on FA, many connections from the frontal pole, right inferior temporal and left occipital regions were highlighted as being predictive of being female (Figure 3a). On the other hand, we saw that many connections from right accumbens, right putamen and left entorhinal were predictive of being male. In general, right thalamus, right caudate, right putamen, left isthmus cingulate and left superior frontal regions were considered by the model as being hubs of important connections for sex classification.

#### Age prediction gradient maps

According to the gradient attribution map for age prediction (AgeGradient) based on SC, many connections from pallidum, temporal pole and medial orbital frontal regions are predictive of being older, while connections from hippocampus, right ventral diencephalon and right transverse temporal regions are predictive of being younger (Figure 2b). In general, the right thalamus, right accumbens areas, right ventral diencephalon, right medial orbital frontal and right precuneus were considered by the model as being hubs of important connections for age prediction.

According to AgeGradient based on FA, many connections from accumbens, isthmus cingulate and right insula regions are predictive of being older, while connections from pars opercularis, pars traingularis, superior frontal and left lateral orbital frontal were predictive of being younger (Figure 3b). In general, the thalamus, putamen, right caudate, left ventral diencephalon and left isthmus cingulate regions were considered by the model as being hubs of important connections for age prediction.

#### *g*-factor prediction gradient maps

It was found that adding covariates to the DL model helped to reduce confounding effects as seen in classical methods. The correlation between gradient attribution map for *g*-factor prediction without covariates (GGradient) and AgeGradient was -0.2722, while correlation between gradient attribution map for *g*-factor prediction with covariates (GCovGradient) and AgeGradient was -0.0996, averaged across cross-validation folds. We hypothesised that the addition of age and sex covariates to the model would have accounted for the effects of these covariates on the *g*-factor and we here therefore report the *g*-factor gradients for the DL model which includes these covariates. We note that according to GCovGradient based on SC, many connections from thalamus, left superior temporal and right parahippocampal regions were chosen as being predictive of higher *g*-factor, while most connections from medial orbital frontal, right amygdala and left lateral orbital frontal regions were predictive of lower *g*-factor (Figure 2c). In general, right thalamus, left putamen, left hippocampus, right superior parietal and right precuneus regions were considered by the model as being hubs of important connections for predicting cognitive *g*-factor (Figure 2c).

Adding covariates had similar effects for the models with FA as what we saw for SC. The correlation between GGradient and AgeGradient was -0.2581 while correlation between GCovGradient and AgeGradient was -0.0852, averaged across folds. According to GCovGradient based on FA, many connections from the left lingual, left rostral anterior cingulate and right entorhinal regions are predictive of higher cognitive *g*-factor, while most connections from left temporal pole, right caudal anterior cingulate and left pallidum regions are predictive of lower cognitive *g*-factor (Figure 3c). In general, ventral diencephalon, right insula, right inferior temporal, right caudate and right isthumus cingulate regions were considered by the model as being hubs of important connections for predicting cognitive *g*-factor.

#### MHQ-factor prediction gradient maps

According to gradient attribution map for MHQ-factor prediction with covariates (MHQCovGradient) based on SC, many connections from right middle temporal, left entorhinal and left superior frontal regions were chosen as being predictive of higher MHQ-factor score (higher MHQ-factor score representing higher psychopathology, while most connections from left pericalcarine, left superior temporal and right caudate regions were predictive of lower MHQ-factor score (Figure 2d). In general, superior temporal, left precuneus, left putamen, right superior parietal and left lingual regions were considered by the model as being hubs of important connections for predicting MHQ-factor score.

According to PCovGradient based on FA, many connections from left amygdala, left lateral orbital frontal and left pars triangularis regions were chosen as being predictive of higher MHQ-factor score, while most connections from left insula, right pallidum and right amygdala regions were predictive of lower MHQ-factor score (Figure 3d). In general, insula, left ventral diencephalon, left posterior cingulate, left middle temporal and right inferior temporal regions were considered by the model as being hubs of important connections for predicting MHQ-factor score.

#### Interpretation of edge importance for each prediction task

In order to verify if small subsets of edges were predominantly responsible for performance at each prediction task, kurtosis measures of the edge-gradient distributions were examined to evaluate whether the probability mass was concentrated around the mean. This way we assessed the gradients for age, *g*-factor and MHQ-factor prediction. It was found that the kurtoses of gradients for age, *g*-factor and MHQ-factor predictions based on SC were respectively 43.67, 10.93, and 11.50 (Figure 4a), and were respectively 19.24, 4.36 and 5.16 based on FA (Figure 4b). This implies that there were more edges having gradients close to the mean (zero) for age prediction compared to the other two prediction tasks. This indicates that the model mainly relied on a smaller but more robust subset of edges for age prediction, but on wider ranges but less robust subset of edges for *g*-factor and MHQ-factor predictions. Figure 4 shows the histograms of gradients for each prediction task based on SC and FA modalities.

**Figure 4:**
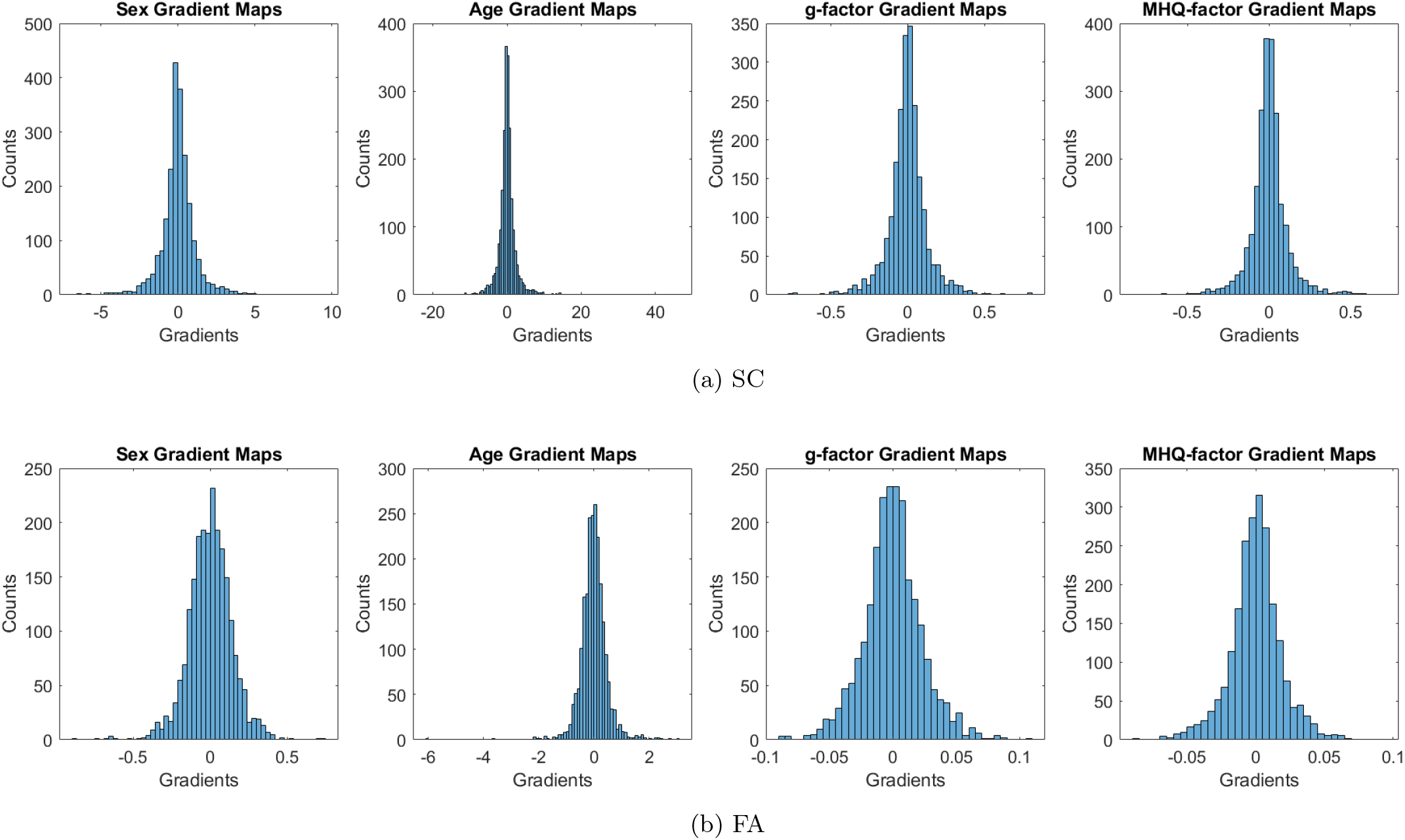
The histograms of gradients based on SC and FA for each prediction task. FA = fraction anisotropy; SC = streamline count. The kurtosis measures of gradient distributions for age, *g*-factor and MHQ-factor predictions based on SC were 43.67, 10.93, and 11.50 respectively, and were 19.24, 4.36 and 5.16 based on FA. This implies that there were more edges having gradients close to the mean (zero) for age predictions than in the other 2 prediction tasks. This indicates that the model relied on a smaller subset of edges for age prediction, but relied on a wide range of edges for *g*-factor and MHQ-factor predictions.

### Comparing prediction performance with classical machine learning approaches

In addition to BainNetCNN, we also applied Ridge Regression, LASSO Regression, linear SVM and Kernel Ridge Regression (KRR) to predict sex, age, *g*-factor and MHQ-factor. Without covariates, all four alternative prediction algorithms (Ridge Regression, LASSO Regression, linear SVM and KRR) performed the best with SC data on most prediction tasks with some exceptions (Ridge Regression and KRR performed best on ISOVF weights for age prediction). When predicting *g*-factor and MHQ-factor with additional covariates, linear methods (Ridge Regression, LASSO Regression, linear SVM) performed better with FA and OD rather than SC data, while KRR performed the best with SC data. The full results and the AUC ROC curves for sex classification are included in section B.3 of the Supplementary Materials. Overall, we found performance of the classical ML methods to be comparable to that of the BrainNetCNN. Tables 3a - 3d show the predictive performance of the five models respectively on sex classification, age prediction, as well as *g*-factor and MHQ-factor predictions (both with age and sex covariates added), based on streamline counts.

**Table 3:** Performances of the five tested algorithms on four different prediction tasks with connectivity measures based on streamline counts. The linear ML models have comparable performance with the non-linear model (KRR) and the deep learning model (BrainNetCNN) on all four evaluated prediction tasks.

| Model | BrainNetCNN | Ridge Logistic Regression | LASSO Logistic Regression | SVM | KRR |
| --- | --- | --- | --- | --- | --- |
| **Validation** | 89.18 (0.70) | 89.67 (1.01) | 88.22 (1.42) | 89.77 (0.91) | 86.44 (1.30) |
| **Training** | 94.10 (1.28) | 93.75 (1.40) | 90.41 (0.29) | 93.90 (0.89) | 88.09 (0.36) |
| **Test** | 86.91 (0.72) | 87.54 (0.48) | 85.90 (0.68) | 87.27 (0.56) | 84.28 (0.36) |

| Model | BrainNetCNN | Ridge Regression | LASSO Regression | SVM-R | KRR |
| --- | --- | --- | --- | --- | --- |
| **Validation** | 4.153 (0.727) | 4.221 (0.713) | 4.238 (0.71) | 4.318 (0.698) | 4.402 (0.685) |
| **Training** | 3.634 (0.795) | 3.773 (0.779) | 3.897 (0.763) | 3.975 (0.738) | 4.271 (0.707) |
| **Test** | 4.245 (0.706) | 4.258 (0.698) | 4.282 (0.695) | 4.357 (0.684) | 4.434 (0.669) |

| Model | BrainNetCNN | Ridge Regression | LASSO Regression | SVM-R | KRR |
| --- | --- | --- | --- | --- | --- |
| **Validation** | 0.264 (0.756) | 0.270 (0.748) | 0.268 (0.749) | 0.270 (0.745) | 0.245 (0.759) |
| **Training** | 0.354 (0.731) | 0.330 (0.735) | 0.335 (0.733) | 0.316 (0.730) | 0.354 (0.728) |
| **Test** | 0.244 (0.769) | 0.248 (0.764) | 0.248 (0.765) | 0.246 (0.763) | 0.257 (0.768) |

| Model | BrainNetCNN | Ridge Regression | LASSO Regression | SVM-R | KRR |
| --- | --- | --- | --- | --- | --- |
| **Validation** | 0.204 (0.758) | 0.223 (0.767) | 0.226 (0.768) | 0.224 (0.794) | 0.207 (0.774) |
| **Training** | 0.335 (0.738) | 0.281 (0.753) | 0.279 (0.755) | 0.286 (0.750) | 0.269 (0.758) |
| **Test** | 0.231 (0.763) | 0.244 (0.772) | 0.237 (0.776) | 0.230 (0.794) | 0.229 (0.780) |

(d) Performance of the five tested algorithms on mental health MHQ-factor prediction (correlation with mean absolute error in brackets) with connectivity measures and common covariates based on streamline counts

For Ridge Regression, LASSO Regression and linear SVM models we additionally extracted feature beta weights in order to compare them with the BrainNetCNN gradient attribution maps. This was not possible for the KRR model due to non-linear transformation of the feature space.

#### Consistency of the Gradients/Betas across folds as well as across different models

Beta weights in classical ML models were generally more stable across folds compared to gradients from the DL models, likely due to the linear nature of the classical approaches. The betas for the ML models from the three classical methods achieved correlations of *>* 0.68 across folds, for all prediction tasks and for each of the network weightings. The betas were generally more stable for sex classification and age prediction compared to prediction of *g*-factor or MHQ-factor. For DL gradients, we saw that the gradients for age and sex prediction were consistent across folds (correlations *>* 0.74). The gradients were however not as consistent for *g*-factor prediction (correlations of 0.48 *−* 0.6). A higher correlation between gradient maps implies a higher consistency in feature ranking between the models across folds, which is why we claimed that the features were more robust age prediction than in *g*-factor and MHQ-factor prediction. Details of across-fold gradient and beta weight correlations are presented in section B.4.1 of the Supplementary Materials.

Regarding the consistency of the DL gradients with classical ML betas, the betas from linear ridge regression were the most consistent with the gradient attribution maps from BrainNetCNN across different prediction tasks. Spearman correlations between the betas and the gradient attribution maps based on SC and FA were 0.66 *−* 0.77 and 0.49 *−* 0.85 respectively for different prediction tasks, see section B.4.2 of the Supplementary Materials.

The top brain regions predictive of *g*-factor and MHQ-factor within BrainNetCNN and within linear Ridge Regression had a high percentage (≥ 60%) of overlap, within both SC and FA modalities (see section B.4.3 in the Supplementary Materials). This implies that the deep learning models and the linear ridge regression models rated features in highly similar ways.

We additionally found that inclusion of covariates in linear models had similar effects on the betas as on the gradients in BrainNetCNN, when both models were based on SC matrices. Without the included covariates, the betas for *g*-factor prediction (GBetas) and the betas for age prediction (AgeBetas) had average correlation of -0.2592. With covariates, the betas for *g*-factor (GCovBetas) and AgeBetas had average correlation of -0.0527.

## Discussion

In the current study, we applied the BrainNetCNN DL model to connectivity matrices derived from six different structural connectivity modalities, MD, FA, SC and three other novel measures OD, ISOVF and ICVF, to predict sex, age, as well as two other clinically relevant behavioural measures, the cognitive *g*-factor and the mental health MHQ-factor using structural connectome data. All connectome processing and network construction steps were performed locally on diffusion and structural MRI data from the UKB. This is one of the largest structural connectome datasets to-date and offers unprecedented statistical power. The BrainNetCNN, which has a more customized design for connectome-based predictive modelling than classical ML models, has novel E2E and E2N layers specifically designed to capture non-linear combinations of brain connections that could be related to complex phenotypes. Taken together, application of BrainNetCNN on a large sample of locally-derived connectomes represents a substantial advantage of our study.

It was found that the best predictions were generally achieved with SC matrices across all models, though these are likely more susceptible to the confounding effects of age and sex, probably via head size. Adding age and sex as external regressors to the BrainNetCNN model helped improve the prediction of both *g*-factor and MHQ-factor, which demonstrates utility of adding external predictors to the BrainNetCNN model. This is consistent with the previous studies, which have shown that adding relevant covariates to CNN models may lead to better predictive performance (Yamamoto et al., 2020). In our study we found that addition of external regressors (i.e. the covariates) had similar effects on the performance of the DL model, as it had on the performance of the classical linear regression models. A recommendation which follows from our results is that researchers should consider adding clinical and demographic covariates to DL models to improve prediction results.

When comparing BrainNetCNN to the classical ML methods, we found similar performance across all prediction tasks. The KRR model did not outperform the other three linear predictive models. This implies that models which simply consider ROI-to-ROI connections individually and linearly can match state-of-the-art machine learning models when applied to structural connectomes. We further found high correlations between gradient maps from DL models and beta maps from linear models, which indicates that BrainNetCNN estimated the connectivity graph node and edge importance weights in similar ways as linear prediction models. Moreover, inclusion of age and sex covariates had similar effects in both BrainNetCNN and linear models. This potentially implies that there is a lack of exploitable non-linearity in brain scans for phenotype prediction, as previously suggested in Schulz et al. (2020). Taken together, these results indicate that most of the variation between phenotypes in weighted structural connectomes is linear rather than non-linear in nature.

In the current study, we found that the models based on SC measures had the best predictive performance compared to other network weightings. This is in line with the results of the previous studies which showed that use of SC measures led to better performance when predicting Autism Spectrum Disorders and paediatric Traumatic Brain Injury (Payabvash et al., 2019; Raji et al., 2020). SC measures might overall be more sensitive to phenotype and psychopathology-related differences in brain connectivity (e.g. (Oestreich et al., 2019)). We note that the entries in SC matrices followed log normal distributions, while entries in other types of connectivity matrices followed normal distributions, which might have been one reason for superior predictive performance with SC matrices. Another possibility, as suggested in Buchanan et al. (2020), could be that the confounding effect of age and sex via head size on SC weightings was larger than on other types of network weightings, which gave a possible explanation to the disappearance of SC’s superiority in predicting *g*-factor and MHQ-factor after adding sex and age to the prediction models.

As expected, we found prediction of *g*-factor (*r* = 0.249) and the MHQ-factor (*r* = 0.234) to be less accurate than prediction of age and sex. We also found that the feature rankings were more robust in sex and age prediction than in *g*-factor and MHQ-factor prediction. Predicting complex cognitive phenotypes from structural neuroimaging is a difficult task. Existing studies conducted on large community samples found that structural neuroimaging measures tend to account for only modest proportions of variance in intelligence (Cox et al., 2019b; Deary et al., 2021). Prediction of case-control status for specific mental illnesses or modelling of mental health continuum has been a similarly difficult task. While some recent studies have had some success in predicting anxiety (Greening and Mitchell, 2015; Wang et al., 2021) and depression symptom severity (Yu et al., 2021) with functional and structural connectomes (achieving correlations *>* 0.24 between the predicted score and the actual score), they generally had relatively small sample sizes with replication yet to be achieved in larger cohorts. Our study extends these results and suggests that brain regions and connections are differentially important for prediction of behavioural and mental health phenotypes such as the *g*-factor and MHQ-factor (Cox et al., 2019b; Dubois et al., 2018).

Our analysis of SC gradient maps revealed the putamen, right precuneus, right thalamus and left superior temporal regions as hubs of important connections for all prediction tasks. For FA gradient maps, we found that the right isthmus cingulate, right ventral diencephalon and right insula were selected as hubs of important connections for all prediction tasks. The relative importance of the regions shown here in the context of prediction modelling should not be interpreted from a functional perspective. In other words, the estimated relative region importance for *g*-factor prediction must not be taken to indicate importance of that region for e.g. the cognitive processes underpinning the *g*-factor. This is because the collinearity among regional brain properties results in the selection of some predictors that carry global information, with other predictors being used for fine tuning of the prediction model. The low-to-medium correlations between the beta coefficients (from ML models) and gradient maps (from DL models) lends some credence to such hypothesis. In-depth interpretations on drawing linkage between structural network weights and behavioural measures will require a different type of analysis technique.

Some limitations of the current study should be mentioned. First, the UKB consists of healthier, highly educated and older individuals (Fry et al., 2017), which may induce bias in model training and the derivation of *g*-factor and MHQ-factor. Second, it is possible that the representation of structural connectivity in the form of adjacency matrices might have limited the deep learning model’s potential to utilise the latent non-linear properties of the brain networks, thus leading to suboptimal performance on prediction of the *g*-factor and MHQ-factor. Another possibility, as described in both He et al. (2020) and Schulz et al. (2020), is that a much larger sample size may be needed for DL models to show superiority over classical ML methods. We note that greater sample sizes will soon be feasible and it will be advantageous for uncovering the relationship between prediction performance and sample size as well as unlocking the potential of DL models. Fourth, BrainNetCNN has novel layers designed for preserving the topology of adjacency matrices. However, the setup of the E2E and E2N layers only allowed aggregating values in the 1-hop neighbourhood (primary neighbours) and this might have limited its ability to explore long-range connections. This could have limited the DL model ability to learn complex non-linear relationships. A simple adjustment could be to alter the network architecture by stacking a number (e.g. *k*) of E2E layers, which would enable the model to explore brain region connections to k-hop rather than 1-hop neighbours. In this case, however, the simpler and shorter-range connections (i.e. 1-hop to (*k −* 1)-hop neighbourhood) may be ignored and not used for prediction in the top layers. To overcome these limitations, future studies could modify the BrainNetCNN architecture according to the idea of residual connections in deep neural networks (He et al., 2016) – which would enable the model to take into account both direct and indirect longer-range connections between brain regions.

The MHQ-factor in this study was derived from online MHQ items collected by the UKB and they were not clinical diagnoses. While it might not be adequate to represent the general dimension of psychiatric disorders, it can be interpreted as a measurement of general mental health that is potentially more informative than binary case control distinctions (Coleman, 2021). We acknowledge that *p*-factor derived from factor modelling is less well validated as in other measures, for example the *g*-factor, and results should be considered accordingly (Cervin et al., 2020; van Bork et al., 2017; Watts et al., 2020). Nonetheless, other studies do support the existence of general dimensions to capture psychopathology (Caspi et al., 2020; Lund et al., 2020; Plana-Ripoll et al., 2019; Sprooten et al., 2022), and further investigate the polygenic *p*-factor derived from polygenic scores for different psychiatric disorders (Allegrini et al., 2020; Selzam et al., 2018). Most current derivations of *p*-factor rely on binary or ordinal data which reduces variance and thus statistical power. Future studies may explore *p*-factor derivations based on continuous variables.

A further, more general limitation of our study is that there is not yet an agreed methodology for constructing structural brain networks from dMRI data (Qi et al., 2015). Connectomic measures are known to be sensitive to the network construction methodology, and different predictive modelling results might be achieved with network construction approaches other than those applied here. dMRI itself only enables noisy and indirect measurement of water molecule diffusion, and thus faithful reconstruction of brain connectivity remains challenging (Buchanan et al., 2020; Jones et al., 2013). These limitations will be addressed in the future with further advances in both structural imaging and tractography methods.

To summarize, we verified that adding relevant external predictors to the BrainNetCNN could potentially aid the prediction. The BrainNetCNN DL model did not perform substantially better compared to classical ML approaches when applied to predict age, sex, *g*-factor or MHQ-factor from connectomic data. Comparison of gradient maps from DL models and beta maps from linear ML models suggested that these methods ranked the features in similar ways. Results also showed that the BrainNetCNN model treated the additional covariates (age and sex) in a similar way as did the classical linear regression models. Overall, results of our study imply that additional model complexity may not improve prediction of complex phenotypes from structural connectomic data.

## Materials and Methods

### Materials

Participants were recruited and brain imaging was completed as part of the UKB study. The UKB field IDs for the cognitive tasks and mental health questionnaire (MHQ) items used in this study are presented in section A.1 of the Supplementary Material. Details of the UKB data can be found at https://biobank.ndph.ox.ac.uk/showcase/label.cgi?id=100000. As a brief summary, four cognitive tasks were chosen for deriving the *g*-factor and 14 MHQ items were chosen for derivation of the MHQ-factor.

#### Participants

A subset of the UKB participants underwent brain MRI at the UKB imaging centre in Cheadle, Manchester, UK. *N* = 9, 858 participants with compatible T1-weighted and dMRI data were collected from the UKB for which we derived connectomes locally, as described below.

#### MRI Acquisition

All imaging data were acquired using a single Siemens Skyra 3T scanner. 3D T1-weighted volumes were acquired using a MP RAGE sequence at 1 *×* 1 *×* 1 mm resolution with 208 *×* 256 *×* 256 field of view (FOV). The dMRI data were acquired using a spin-echo EPI sequence (50 *b* = 1000*s/mm*^2^, 50 *b* = 2000*s/mm*^2^ and 10 *b* = 0*s/mm*^2^) resulting in 100 distinct diffusion-encoding directions, FOV = 104 *×* 104 mm, imaging matrix = 52 *×* 52, 72 slices, slice thickness = 2 mm. *N* = 831 (∼ 8.4% of total *N*) participants with missing dMRI data or processing failure were excluded from the analysis. Details of the MRI protocol and imaging data processing can be found in (Alfaro-Almagro et al., 2018; Miller et al., 2016).

#### Network Construction

The network data and methods of network construction used in this study have been published previously (Buchanan et al., 2020; Madole et al., 2021) and are outlined below. All network construction steps were performed locally on MRI data. Each T1-weighted image was segmented into 85 distinct neuroanatomical Regions-Of-Interest (ROI) using volumetric segmentation and cortical reconstruction (FreeSurfer v5.3.0), 34 cortical structures per hemisphere were identified using the Desikan-Killany atlas (Desikan et al., 2006). Brain stem, accumbens area, amygdala, caudate nucleus, hippocampus, pallidum, putamen, thalamus and ventral diencephalon were also extracted with FreeSurfer. Outputs were visually inspected and those which failed to meet quality control (QC) standards were removed (Cox et al., 2019a). *N* = 842 (∼ 8.5% of total *N*) participants with incomplete FreeSurfer output or processing failure were excluded from the analysis. A cross-modal nonlinear registration method was used to align ROIs from T1-weighted volume to diffusion space (skull stripping (Smith, 2002), initial alignment by affine transformation with 12 degrees of freedom (FLIRT; (Jenkinson and Smith, 2001)) followed by a nonlinear deformation method (FNIRT; (Andersson et al., 2007))).

Networks were constructed by identifying connections between all ROI pairs. The endpoint of a streamline was recorded as the first ROI encountered when tracking from the seed location. Successful connections were recorded in an 85 *×* 85 adjacency matrix. In total, six network weightings, FA, MD, SC, OD, ISOVF and ICVF, were computed. For each weighting, an adjacency matrix was computed with each element, *a*_*ij*_, recording the mean value of the diffusion parameter in voxels identified along all interconnecting streamlines between nodes *i* and *j*. All matrices were made symmetric since afferent and efferent connections are indistinguishable for tractography. Self-connections were removed, setting diagonal entries to zero (Buchanan et al., 2020). *N* = 2 participants with disconnected connectivity matrices were excluded.

In total, 8, 183 participants (45.1–78.5 years of age, 3,869 male) remained after participants were excluded at QC stage or due to failure in processing. Although it was reported in Madole et al. (2021) that *N* = 157 participants (< 2%) might have dementias and neurological syndromes, for example stroke and brain injuries, they found that excluding these participants from the sample did not result in significant differences in their primary outcome measures (Madole et al., 2021). We therefore kept the full sample for the following analysis. On average, 6.01 million streamlines were seeded per subject of which 1.49 million (24.9%) were found to successfully connect between nodes following the tracking procedure and removal of self-connections. For each participant, networks were produced from the same set of streamlines, where the range of values for MD is 0 - 0.003 *×*10^*−*3^*mm*^2^*/s*, for FA is 0 - 0.9, for SC is 0 - 4.03 *×*10^4^, for OD is 0 - 0.9, for ISOVF is 0 - 1 and for ICVF is 0 - 1. Before any thresholding was introduced, the mean value of network density (percentage of non-zero entries in an adjacency matrix) across subjects was 68.1% (SD = 3.1). Proportional-thresholding was used to keep only connections present in at least 2/3 of subjects, which resulted in connection density of ∼60% after thresholding.

#### Cognitive Tasks and *g*-factor

The cognitive tasks included the Verbal Numerical Reasoning (VNR), Reaction Time (RT, log-transformed), Pairs Matching (Pairs Match, log (x+1) transformed) and Prospective Memory (Fawns-Ritchie and Deary, 2020; Lyall et al., 2016) tests. Table 4 shows participants characteristics, demographic information and cognitive task scores. Missing values were imputed in R with the missMDA package (Josse et al., 2016). Description of the cognitive tasks’ scores is presented in section A.1.1 of the Supplementary Materials.

**Table 4:**
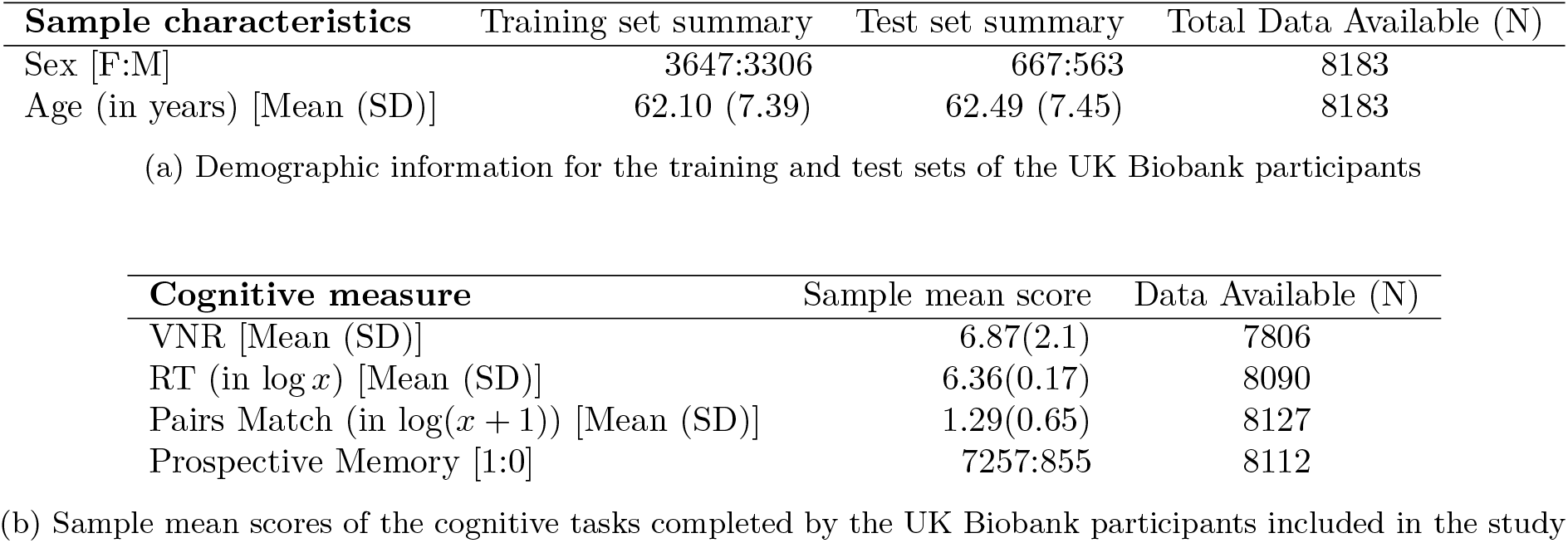
Summary characteristics of the UK Biobank participants included in the study. VNR: Verbal Numerical Reasoning. RT: Reaction Time. Pairs Match: Pairs Matching. For the prospective memory test, **1** means recall at the first attempt and **0** otherwise.

Principal component analysis (PCA) implemented in *pcacov* function in MATLAB 2020a was applied to the 4 cognitive tasks as an intermediate step to derive the cognitive *g*-factor value for each participant. The principal component score, *g*-factor, was computed as the sum of *z*-normalised cognitive task scores multiplied by the respective first unrotated component coefficients from *pcacov*. The *g*-factor explained 34.55% of the variance. To avoid data leakage, the principal component coefficients were computed with exclusion of the test data (see below). Excluding the missing values and test data in the cognitive tasks, 6558 samples were available for computing the principal component coefficient. The *g*-factor was *z*-normalised based on the training data. The resulting *g*-factor had a range [-4.3661, 2.7664]. The standardised loadings of scores for each task on the *g*-factor are shown in Table 5.

**Table 5:** Standardised loadings of individual cognitive test scores on the cognitive *g*-factor. VNR: Verbal Numerical Reasoning. RT: Reaction Time. Pairs Match: Pairs Matching. PropVar: Proportion of variance explained by the *g*-factor.

| **Cognitive Tasks** | **Loadings** |
| --- | --- |
| VNR | 0.6914 |
| RT | -0.5514 |
| Pairs Match | -0.5186 |
| Prospective Memory | 0.5752 |
| PropVar | 0.3455 |

#### MHQ-factor

The derivation of the general mental health score, the MHQ-factor, followed the same principles as described in (Lund et al., 2020). Lund et al. (2020) applied principal component analysis (PCA) to the 14 mental health questionnaire (MHQ) items from the UKB which measured common forms of depression, anxiety, psychotic experiences and substance abuse, and took the first unrotated principal component score as the MHQ-factor. In this study, the same set of MHQ items was used to derive the MHQ-factor. Out of 8,183 participants, 6,247 participants had fewer than 3 missing values on these 14 MHQ items. The missing values were imputed in R with the missMDA package (Josse et al., 2016).

Since all MHQ items were categorical variables, the polychoric correlation was more appropriate than Pearson’s correlation for application of PCA. We applied PCA on polychoric correlation of the 14 items and took the first unrotated principal component score as the MHQ-factor. A comparison of the two PCA methods (using polychoric and Pearson correlation) is presented in section A.2.1 of the Supplementary Materials. We checked the variance explained, factor congruence between the two methods, and assessed correlations between the principal component scores derived using the two methods. This motivated selection of polychoric correlation for the PCA. The derived MHQ-factor explained 49.60% for the variance. Figure 5 shows the loadings of the individual MHQ items on the principal components. We saw strong positive loadings of depressive/anxiety items on the first PC and positive loadings of psychotic experiences items on the second PC, which is similar to the results reported in (Lund et al., 2020). It was also found that the MHQ-factor was not significantly associated with the *g*-factor (Pearson’s Correlation = 0.0025, *p*-value = 0.8483).

**Figure 5:**
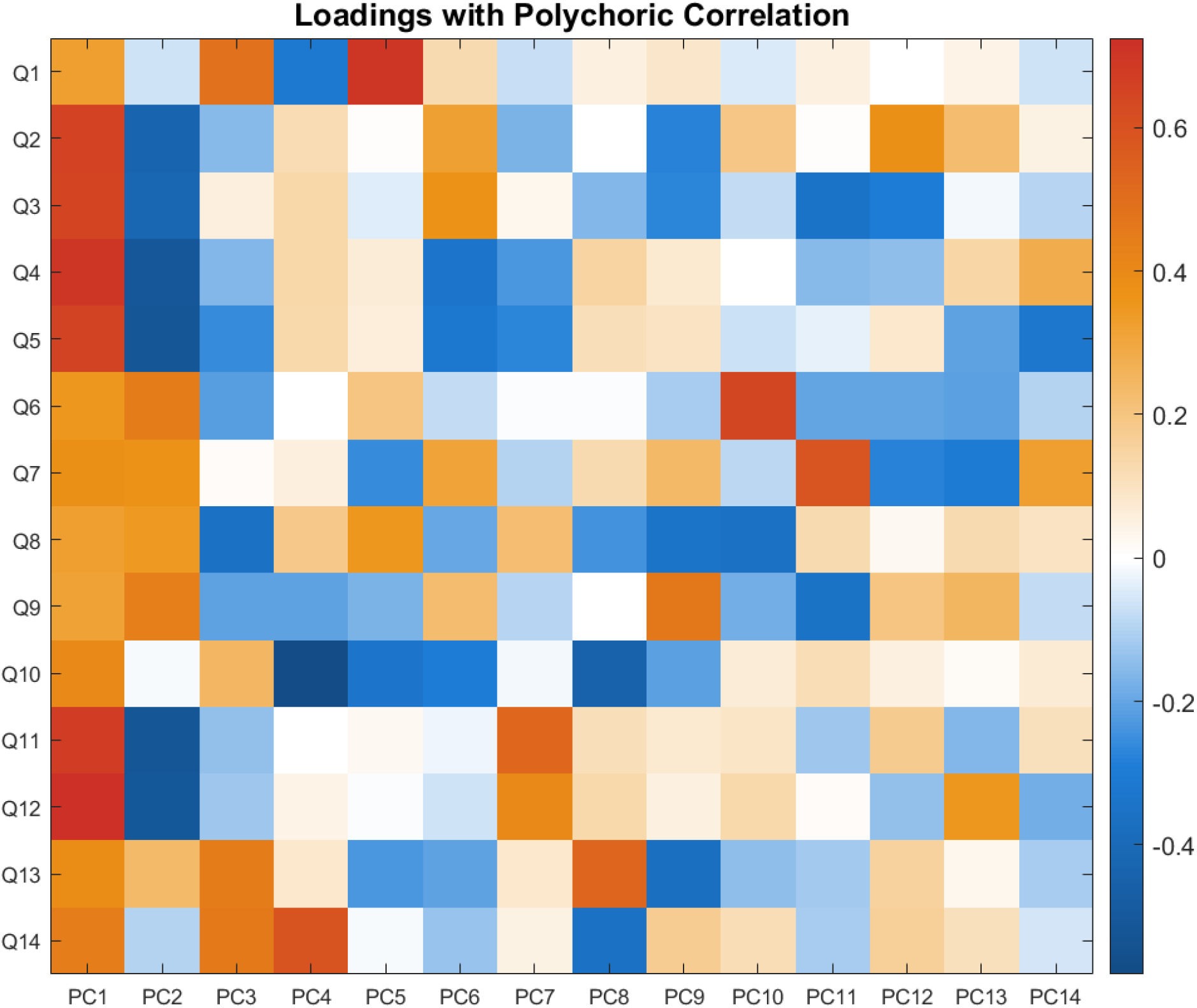
Loadings of the 14 MHQ items in UKB on the principal components derived with PCA. We saw strong positive loadings of depression/anxiety items on the first PC and positive loadings of psychotic experience items on the second PC. Please see Supplementary Table 1 in section A.2.2 for exact values of standardised item loadings on principal components. Q1: Ever addicted to any substance or behaviour ; Q2: Ever felt worried, tense, or anxious for most of a month or longer; Q3: Ever worried more than most people would in similar situation ; Q4: Ever had prolonged loss of interest in normal activities ; Q5: Ever had prolonged feelings of sadness or depression ; Q6: Ever heard an un-real voice ; Q7: Ever believed in an un-real conspiracy against self ; Q8: Ever seen an un-real vision ; Q9: Ever believed in un-real communications or signs; Q10: Ever self-harmed ; Q11: Ever sought or received professional help for mental distress ; Q12: Ever suffered mental distress preventing usual activities ; Q13: Ever had period of mania / excitability ; Q14: Ever had period extreme irritability

### Methods

#### BrainNetCNN Components

We chose the BrainNetCNN model as it is specifically designed for connectomic data. While additional transformation to the graph data is often needed for other methods (e.g. mapping connectomic data to participant-wise similarity matrix using graph kernels or to *n*-dimensional Euclidean space using graph embedding), the information aggregation process within the custom layers within the BrainNetCNN model preserves the topological locality of connectivity adjacency matrices. Kawahara et al. (2017) provide clear formulations of the special layers for the BrainNetCNN (Kawahara et al., 2017). Briefly, the layers user in the network are following:

1. Edge-to-Edge (E2E) layer: This is a convolution layer with a cross-shaped filter where the (*i, j*) entry output is given by the weighted sum of the *i*-th row and weighted sum of the *j*-th column. This can be written as:

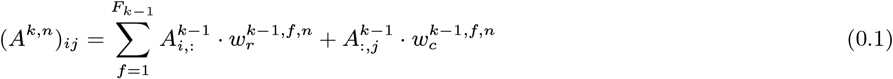

where *A*^*k,n*^ is the filtered adjacency matrix, *A*_*i*,:_ is the *i*-th row of *A* and *A*_:,*j*_ is the *j*-th column of *A*, and 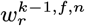 and 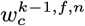 are the learnt row and column weights for the *n*-th filter at the *k −* 1-th layer, respectively. *F*_*k−*1_ is the number of feature maps at the *k −* 1-th layer.
2. Edge-to-Node (E2N) layer: An E2N layer is equivalent to adding a spatial 1D convolutional row filter of the adjacency matrix to a transposed spatial 1D convolutional column filter of the adjacency matrix. Kawahara et al. (2017), showed that combining the row and column filters does not result in improvement of model performance when compared to using only the row filter, and hence only the row filter was applied in the current study.

#### Network Architecture

##### Architecture without covariates

The architecture of the deep neural network is presented in Figure 6a. This is a simpler version of the original architecture described in Kawahara et al. (2017). We were able to significantly reduce the number of layers and filters while maintaining prediction performance. Moreover, either increasing the number of filters per layer or the number of hidden layers made the overfitting more severe. We have also experimented with hyperparameter tuning for the E2E, E2N and N2G layers, aiming to optimise scales for Leaky Rectified Linear Unit and the dropout rate using Bayesian Optimisation. The hyperparameter optimisation in BrainNetCNN was performed on a fixed predefined validation set (rather than through cross-validation) to enable reasonable optimization times. The accuracies were similar to those without tuning and we therefore proceeded with fixed hyperparameters (Figure 6) in our further investigations. Full results with hyperparameter tuning are shown in the Supplementary Materials section B.2.

**Figure 6:**
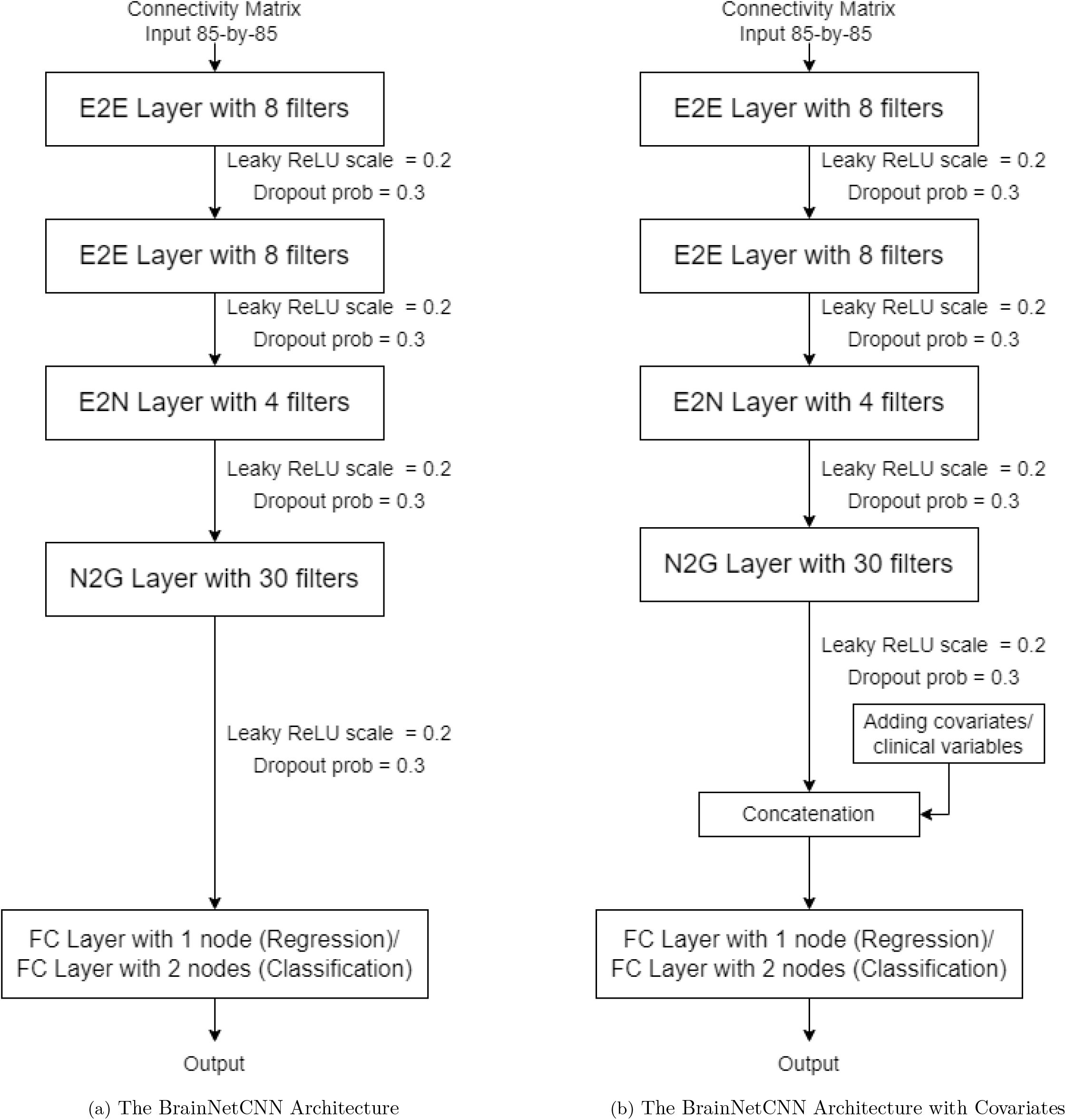
Two BrainNetCNN Architectures. E2E: Edge-to-Edge, E2N: Edge-to-Node, N2G: Node-to-Graph, FC: Fully connected, Leaky ReLU: Leaky Rectified Linear Unit, prob: probability

##### Architecture with covariates

The *g*-factor and MHQ-factor are known to be significantly associated with age and sex, and hence consideration of these factors in the predictive model can improve its performance. To take into account age and sex covariates, the model was update with additional layers. Specifically, a concatenation layer was connected to the N2G layer to include the covariates. Figure 6b shows the architecture of the updated model.

The loss function for sex classification was cross entropy and the loss function for the other three prediction tasks was mean square error.

#### Network Input

The SC matrices were first made symmetric by addition of the transpose and division by 2. We then performed maximum value normalisation on the symmetric SC matrices. The maximum value normalisation was performed on all modalities as we previously found this to be the most effective normalisation (Yeung et al., 2020). Maximum value normalisation is formulated as follows:

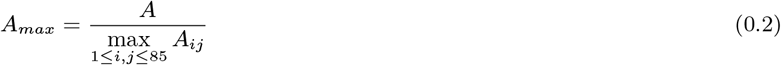

Since it is known that the head size has great influence on the streamline counts, we computed the correlation between the intracranial volume (ICV) and the total sum of entries in the SC matrices before and after normalisation. We found that the correlation was 0.7405 (*β* = 0.7988, *p <* 1 *×* 10^*−*324^) before normalisation and -0.0701 (*β* = *−*1.023 *×* 10^*−*5^, *p* = 3.784 *×* 10^*−*11^) after normalisation. Therefore, we saw that the maximum value normalisation was able to largely remove the effect of ICV. Figure 7 shows the mean connectivity matrices computed across all participants for different weightings and histograms of edge weights pooled across participants. The SC weights were shown in log-scale. The five network weightings except for SC had similar distribution shapes. MD, FA, OD and ICVF had skewness in the range [-0.3, 0.3]. ISOVF had a skewness of 1.36 and SC has a skewness of 8.01.

**Figure 7:**
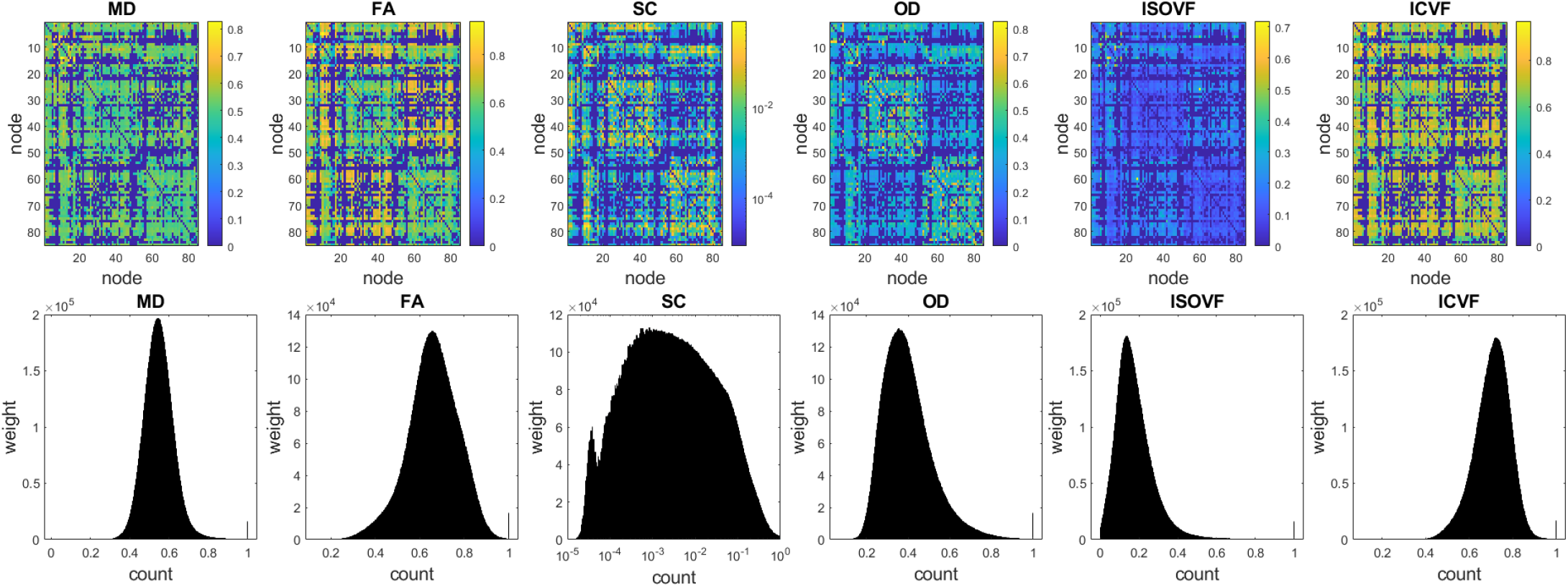
Top: 85 *times* 85 mean connectivity matrices of inter-region connection weights averaged across all participants (N = 8,183) for six network weightings after performing maximum value normalisation. The two blocks along the diagonal in each case correspond to the left and right hemispheres. Bottom: the corresponding histograms of nonzero edge weights pooled across all participants for each weighting (SC is log-scaled). MD = mean diffusivity; FA = fraction anisotropy; SC = streamline count; OD = orientation dispersion; ISOVF = Isotropic volume fraction; ICVF = Intracellular volume fraction. The SC weights were shown in log-scale. Apart from the SC weights, the other five network weightings had similar distribution shape. MD, FA, OD and ICVF had skewness in the range [-0.3, 0.3]. ISOVF had a skewness of 1.36 and SC has a skewness of 8.01.

#### Gradient Attribution Map

Gradient Attribution Map were computed to check which connectivity matrix entries influenced prediction the most (Simonyan et al., 2013). This is similar to computing the partial derivative of the class score with respect to each entry of the *A*^*n*^ matrix, where the magnitudes of the derivatives represent the degrees of influence on the final class score, *y*^*n*^, for some participant n. This means that the magnitudes of gradients approximately tell us how the model ranks the structural connectome feature importance with respect to the evaluated prediction task. The sign of the gradient indicates whether the connectome feature is positively or negatively predictive of the target variable. Gradients thus partly inform us of the model’s strategy in making its predictive decisions. By computing correlations between gradient maps for different models (i.e. trained for different prediction tasks), it is thus possible to assess similarity in the model strategies for completing these tasks.

Due to the stochastic nature of the neural network training process, the gradient maps may be different across different training runs and across different parameter initialisations. In our study the BrainNetCNN models were therefore trained 20 times for each cross-validation (CV) partition with randomised parameter initialisations. Since we used 5-fold cross validation, there were 100 BrainNetCNN models trained in total. We then took the average of the gradient maps from the 100 models for each modality and each prediction task.

Since *A*^*n*^ is symmetric, we expected that 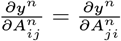. However, in reality, since the column weights and row weights within the filters were trained independently, it was found that not only 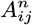 and 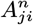 had different magnitudes, but some of the pairs even had opposing signs. To solve this issue, we summed each gradient map with its transpose, thus getting a single gradient estimate for each ROI-to-ROI connection.

Predictive power of each node (ROI) was defined as the sum of the gradients of the node’s edges. This enabled determining whether the node was generally predictive of higher or lower outcome values. Importance of each node was defined as the sum of the absolute values of the node edges’ gradients This was used to determine whether a node was a hub of important connections for prediction. We report the top regions relevant for each prediction task based on these two measures (predictive power and importance) in section B.4.3 of the Supplementary Materials. We should note that our assessment of node importance was purely based on the magnitudes and directions of the gradient map entries. We thus do not claim that the regions defined as important for prediction in this study were the only ones related to (or responsible for) the evaluated phenotypes.

#### Model Hyperparameters

For both models we used Adam optimiser (Kingma and Ba, 2014) for weight updates, with the same set of hyperparameters used in training: learning rate = 0.001, gradient decay factor = 0.9, squared gradient decay factor = 0.9, mini batch size = 128, validation frequency = 50, training epoch = 200. The training stopping points corresponded to the best validation accuracy.

#### BrainNetCNN Validation and Testing

The participants were first sorted according to their participant ID in ascending order. The test set consisted of the first 15% of the total sample (∼ 1230 participants for prediction of sex, age and cognitive functioning, ∼ 940 participants for MHQ-factor – a slightly smaller number because only 6,247 participants had both structural connectomes data and responses to MHQ items available). 5-fold CV was performed on the rest of the data. The same set of CV fold splits was used across the imaging modalities to enable direct comparisons. In each iteration of the 5-fold CV, a BrainNetCNN model was trained on the corresponding training fold for 200 epochs and the deep learning weights at each epoch were recorded. This resulted in 200 sets of weights representing different phase of learning within the training process. Each set of weights (or epoch weights) gave a corresponding validation accuracy/correlation, and the set of weights that yielded the best validation accuracy were taken as the optimal weights for the iteration and were used for performance evaluation on the test data. Test accuracies were averaged across the folds to get performance estimates.

#### Classical Machine Learning Approaches

We compared the DL models with linear versions of three common classical methods: Ridge Regression, LASSO Regression, and linear SVM. The beta weights can be efficiently extracted from these three types of models and therefore enabled comparison with the BrainNetCNN gradient attribution maps. We also applied Kernel Ridge Regression (KRR). Multiple studies suggest that KRR may have comparable performances with DL methods (He et al., 2020; Mihalik et al., 2019; Schulz et al., 2020). Input features to these models were the individual ROI-to-ROI connections specified in the normalised adjacency matrices (i.e. the non-zero upper/lower triangular entries of the matrices).

In addition to the predictive analyses based on the connectomic data we also tested using only age and sex to predict the *g*-factor and MHQ-factor. The models were underfit and we therefore did not examine them further.

##### Ridge Regression

Ridge regression is very similar to ordinary least squares regression except for the fact that it has an extra regularisation term:

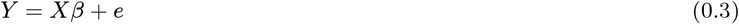

where *Y* is the response variable, *X* is the input feature vectors, *β* are the learnt feature weights, subjected to the following cost function,

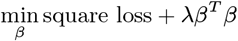

where square loss = (*Y − Xβ*)^*T*^ (*Y − Xβ*). The *L*_2_ regularisation term (*λβ*^*T*^ *β*) is added to prevent overfitting. For logistic regression, the equation is the following,

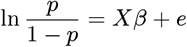

where *p* = *P* (*Y* = 1), and square loss is replaced by logistic loss:

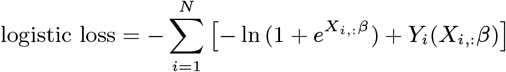

where *N* is the number of observation, *X*_*i*,:_ is the feature vector for observation *i, Y*_*i*_ is the response for observation *i*.

##### LASSO Regression

LASSO regression is similar to Ridge regression except that the L2 regularisation term is replaced by L1 regularisation term, with the same regression equation, but subject to a different constraint:

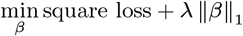

Similarly to logistic Ridge regression, square loss is replaced by logistic loss in logistic LASSO regression.

##### Support Vector Machine

SVM is one of the most popular ML techniques applied in the neuroimaging literature. For classification, SVM constructs hyperplanes between two classes such that the distance between them is as large as possible. The larger the hyperplane margin between classes, the lower the out-of-sample error. SVM regression (SVM-R) is similar to the “soft margin” concept in SVM classification. More details can be found in (Boser et al., 1992) for support vector classification and (Drucker et al., 1997; Ho and Lin, 2012; Vapnik et al., 1997) for support vector regression. Linear kernel was used in this study. The epsilon parameter for linear SVM-R was estimated using the interquartile range of response variable Y.

##### Kernel Ridge Regression

Kernel regression is a non-parametric ML algorithm (Murphy, 2012). Instead of taking the original features as predictors, it takes the subject pairwise-similarities as the predictors for the ML model. Let *X*_*train*_ = {*x*_1_, *x*_2_, *…, x*_*n*_} be the set of feature vectors and *Y*_*train*_ = {*y*_1_, *y*_2_, *…, y*_*n*_} be the set of responses in the training set. For a test subject *x*^*′*^, *y*^*′*^ is estimated through the following equation:

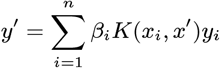

where the *K*(*x*_*i*_, *x*^*′*^) is the similarity between *x*^*′*^ and each feature vector *x*_*i*_ in the training set. *β*_*i*_ are the learnt feature weights, subjected to the cost function similar to the one stated in the section of *Ridge Regression*. With this setup, the dimension is basically equal to the number of observations in the training set in each fold. Similar to He et al. (2020), the similarity *K*(*·, ·*) was chosen to be the Pearson’s correlation between the upper triangular entries of matrices (He et al., 2020).

##### Classical ML Methods Validation and Testing

LASSO regression, Ridge regression and linear SVM methods have single regularisation hyperparameter *λ*. Search range for optimal *λ* consisted of 300 different values ranging from 1e-6 to 1e-1 equally spaced on the logarithmic scale. The validation process for the classical machine learning models was similar to that used with BrainNetCNN. After separating the test set from the rest of the data, a 5-fold CV was performed on the rest of the data. In each iteration of the 5-fold CV, a model was trained on the training fold data with each of the 300 *λ* values, resulting in 300 models. The *λ*_*i*_ that optimized the validation accuracy/correlation was chosen for that iteration, so the *λ*s are different for each fold, each model and each connectivity weight.”

## Data Availability

The UK Biobank's Access Procedures stipulate that participant data can only be made available to approved researchers. Therefore, the data used in this study cannot be made available for public access.

## Acknowledgements

This study was supported by Wellcome Trust awards (References 104036/Z/14/Z; 220857/Z/20/Z), and was also supported by National Institutes of Health (NIH) research grant R01AG054628 which supported CRB, EMT, MEB and SRC. The research was conducted using the UK Biobank resource, with approved project number 10279. Structural brain imaging data from UK Biobank was processed using facilities within the Lothian Birth Cohort group at the University of Edinburgh, which is supported by Age UK (as The Disconnected Mind project), the Medical Research Council (MR/R024065/1), and the University of Edinburgh. This work has made use of the resources provided by the Edinburgh Compute and Data Facility (ECDF). The Population Research Center (PRC) and Center on Aging and Population Sciences (CAPS) at The University of Texas at Austin are supported by National Institutes of Health (NIH) grants P2CHD042849 and P30AG066614, respectively. KMS was supported by Health Data Research UK, an initiative funded by UK Research and Innovation Councils, NIH Research (England) and the UK devolved administrations, and leading medical research charities. SRC was also supported by a Sir Henry Dale Fellowship jointly funded by the Wellcome Trust and the Royal Society (221890/Z/20/Z). AMM and HCW are additionally supported by a UKRI award (Reference MC_PC_17209).

