## Supplementary material for "Predicting sex, age, general cognition and mental health with machine learning on brain structural connectomes": NA

Supplementary Materials

Hon Wah Yeung[1], Aleks Stolicyn[1], Colin R. Buchanan[2,9,10], Elliot M. Tucker-Drob[3,4], Mark E. Bastin[5,9,10], Saturnino Luz[6], Andrew M. McIntosh[1,7], Heather C. Whalley[1,*], Simon R. Cox[2,9,10,*], and Keith Smith[8,*]

[1]Department of Psychiatry, University of Edinburgh, Edinburgh, United Kingdom
[2]Department of Psychology, University of Edinburgh, Edinburgh, United Kingdom
[3]Department of Psychology, University of Texas, Austin, TX, USA
[4]Population Research Center and Center on Aging and Population Sciences, University of Texas at Austin, TX, USA
[5]Centre for Clinical Brain Science, University of Edinburgh, Edinburgh, United Kingdom
[6]Usher Institute, Edinburgh Medical School, The University of Edinburgh, Edinburgh, United Kingdom
[7]Centre for Genomic and Experimental Medicine, Institute of Genetics and Molecular Medicine, University of Edinburgh, Edinburgh, UK
[8]Department of Physics and Mathematics, Nottingham Trent University, Nottingham, United Kingdom
[9]Lothian Birth Cohorts, University of Edinburgh, Edinburgh, United Kingdom
[10]Scottish Imaging Network, A Platform for Scientific Excellence Collaboration (SINAPSE), Edinburgh, United Kingdom
[*]These authors share joint senior authorship

November 22, 2022

### A Materials and Methods

#### A.1 UK Biobank Field ID

The UK Biobank field IDs of the cognitive tasks and mental health questionnaire items used in this study were shown below.

##### A.1.1 Cognitive tasks

**Verbal Numerical Reasoning (UKB Field ID: 20016.2.0)**:

Participants were asked to answer 13 multiple-choice questions and the score is the number of questions answered correctly in two minutes.

**Reaction Time Task (UKB Field ID: 20023.2.0)**:

Two cards were shown on the screen in front of the participants in each trial. They were asked to press the button-box as quickly as possible when the two cards have matching symbols. There were 12 trials and the first 5 were for training. 4 out of the remaining 7 trials have matching cards. The score is the mean time to to press the button-box for these 4 trials.

**Pairs Matching (UKB Field ID: 399.2.2)**:

This task aims at testing the participants' visual memory and the scores were obtained from the second trial. Participants were asked to memorise the patterns 6 pairs of cards. The cards were then faced down and participants needed to correctly identify the pairs of cards with matching patterns. The score was the number of errors made before matching all pairs.

**Prospective Memory (UKB Field ID: 20018.2.0)**:

At the start of the UK Biobank cognitive test battery, participants would see this instruction on the screen "At the end of the games we will show you four coloured symbols and ask you to touch the Blue Square. However, to test your memory, we want you to actually touch the Orange Circle instead.". After completing the cognitive tasks, participants were asked to touch the Blue Square. Participants who correctly touched the orange circle on first attempt were coded 1 and 0 otherwise in this study.

##### A.1.2 Mental Health Questionnaire

| Field ID | Description |
|---|---|
| 20401.0.0 | Ever addicted to any substance or behaviour |
| 20421.0.0 | Ever felt worried, tense, or anxious for most of a month or longer |
| 20425.0.0 | Ever worried more than most people would in similar situation |
| 20441.0.0 | Ever had prolonged loss of interest in normal activities |
| 20446.0.0 | Ever had prolonged feelings of sadness or depression |
| 20463.0.0 | Ever heard an un-real voice |
| 20468.0.0 | Ever believed in an un-real conspiracy against self |
| 20471.0.0 | Ever seen an un-real vision |
| 20474.0.0 | Ever believed in un-real communications or signs |
| 20480.0.0 | Ever self-harmed |
| 20499.0.0 | Ever sought or received professional help for mental distress |
| 20500.0.0 | Ever suffered mental distress preventing usual activities |
| 20501.0.0 | Ever had period of mania / excitability |
| 20502.0.0 | Ever had period extreme irritability |

#### A.2 Principal Component Analysis

##### A.2.1 Comparison between using Pearson's and polychoric correlation

Figure 1 showing the scree plot. For our dataset, the MHQ-factor derived with Pearson's correlation explained 28.03% for the variance, while the MHQ-factor derived with polychoric correlation explained 49.60% for the variance. Th scree plots for both methods indicated that the significant dimensions were spanned by the first 3 eigenvectors, where the eigenvalues were larger than one. The congruence coefficients for the loadings between the two methods of the first three principal components (PC) were 0.9428, 0.9406 and 0.9686 respectively, and the correlation for the first three principal component scores between the two methods were 0.9748, 0.8866 and 0.9738 respectively. As such, it is clear that both methods for deriving the MHQ-factor convey highly similar information. As such, we present results based on polychoric correlation method for the MHQ-factor in all subsequent analyses, as it explained a significantly larger variance.

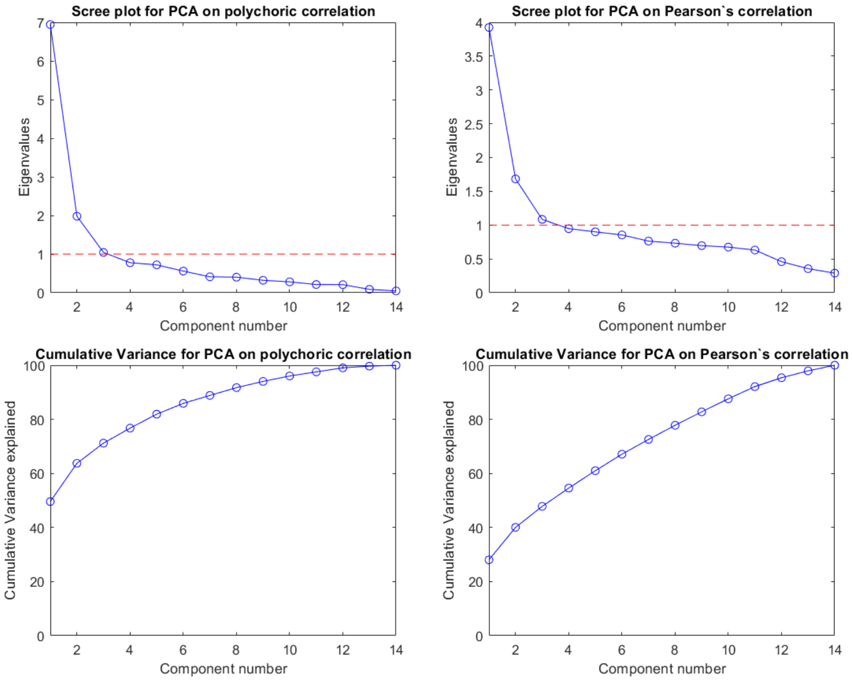


Figure 1: The scree plots and cumulative variance explained for PCA with Pearson's correlation and PCA with polychoric correlation respectively

##### A.2.2 Numeric loadings for PCA with polychoric correlation

Table 1: The principal component loadings on the Mental Health Questionnaire items

| Loadings | PC1 | PC2 | PC3 | PC4 | PC5 | PC6 | PC7 | PC8 | PC9 | PC10 | PC11 | PC12 | PC13 | PC14 |
|---|---|---|---|---|---|---|---|---|---|---|---|---|---|---|
| 20401.0.0 | 0.500 | 0.001 | 0.489 | -0.281 | 0.638 | 0.102 | -0.060 | 0.041 | 0.078 | -0.033 | 0.038 | 0.005 | 0.003 | -0.009 |
| 20421.0.0 | 0.789 | -0.292 | -0.154 | 0.056 | -0.005 | 0.322 | -0.194 | -0.002 | -0.183 | 0.129 | 0.045 | 0.258 | 0.075 | 0.019 |
| 20425.0.0 | 0.787 | -0.294 | 0.086 | 0.065 | -0.078 | 0.353 | -0.013 | -0.143 | -0.171 | -0.131 | -0.161 | -0.237 | -0.047 | -0.021 |
| 20441.0.0 | 0.815 | -0.360 | -0.106 | 0.062 | 0.052 | -0.286 | -0.225 | 0.099 | 0.063 | -0.005 | -0.070 | -0.132 | 0.064 | 0.126 |
| 20446.0.0 | 0.799 | -0.376 | -0.243 | 0.053 | 0.028 | -0.235 | -0.235 | 0.028 | 0.087 | -0.085 | 0.031 | 0.060 | -0.119 | -0.116 |
| 20463.0.0 | 0.616 | 0.613 | -0.205 | 0.014 | 0.162 | -0.058 | -0.016 | 0.014 | -0.096 | 0.377 | -0.076 | -0.109 | -0.062 | -0.020 |
| 20468.0.0 | 0.762 | 0.432 | 0.023 | 0.064 | -0.199 | 0.219 | -0.064 | 0.072 | 0.138 | -0.047 | 0.308 | -0.076 | -0.083 | 0.049 |
| 20471.0.0 | 0.569 | 0.496 | -0.367 | 0.175 | 0.300 | -0.146 | 0.145 | -0.169 | -0.188 | -0.247 | 0.063 | 0.041 | 0.037 | 0.016 |
| 20474.0.0 | 0.632 | 0.565 | -0.202 | -0.204 | -0.126 | 0.151 | -0.049 | -0.015 | 0.301 | -0.094 | -0.216 | 0.069 | 0.077 | -0.018 |
| 20480.0.0 | 0.639 | 0.040 | 0.221 | -0.539 | -0.280 | -0.233 | -0.012 | -0.315 | -0.113 | 0.030 | 0.058 | 0.030 | -0.001 | 0.011 |
| 20499.0.0 | 0.785 | -0.383 | -0.097 | -0.080 | 0.023 | 0.016 | 0.404 | 0.078 | 0.076 | 0.028 | -0.085 | 0.129 | -0.130 | 0.061 |
| 20500.0.0 | 0.846 | -0.336 | -0.093 | -0.033 | -0.045 | -0.027 | 0.269 | 0.103 | 0.058 | 0.079 | 0.137 | -0.123 | 0.164 | -0.076 |
| 20501.0.0 | 0.629 | 0.381 | 0.423 | 0.061 | -0.209 | -0.141 | 0.016 | 0.384 | -0.210 | -0.099 | -0.079 | 0.063 | 0.008 | -0.021 |
| 20502.0.0 | 0.580 | 0.049 | 0.484 | 0.552 | -0.047 | -0.100 | 0.043 | -0.269 | 0.147 | 0.083 | -0.034 | 0.071 | 0.020 | -0.007 |

### B Results from deep learning and classical machine learning methods

#### B.1 Abbreviations in the circular plots

| Abbreviation | Full Name | Abbreviation | Full Name |
|---|---|---|---|
| ctx | Cortex | lingual | lingual |
| lh | left hemisphere | mOFC | medial orbito frontal |
| rh | right hemisphere | MTG | middle temporal |
| THA | thalamus proper | ParaHG | parahippocampal |
| CAU | caudate | ParaCL | paracentral |
| PUT | putamen | ParsOp | pars opercularis |
| PAL | pallidum | ParsOrb | pars orbitalis |
| brain.stem | brain stem | ParsTr | pars triangularis |
| HIP | hippocampus | PCalCC | perical carine |
| AMYG | amygdala | PoCG | postcentral |
| Accumbens | accumbens area | PCC | posterior cingulate |
| VentralDC | ventral diencephalon | PrCG | precentral |
| banksSTS | banks of superior temporal sulcus | precuneus | precuneus |
| cACC | caudal anterior cingulate | rACC | rostral anterior cingulate |
| cMFG | caudal middle frontal | rMFG | rostral middle frontal |
| cuneus | cuneus | SFG | superior frontal |
| entorhinal | entorhinal | SPL | superior parietal |
| fusiform | fusiform | STG | superior temporal |
| IPL | inferior parietal | SMG | supramarginal |
| ITG | inferior temporal | FPC | frontal pole |
| ICgG | isthmus cingulate | TPC | temporal pole |
| lOC | lateral occipital | TTG | transverse temporal |
| lOFC | lateral orbito frontal | insula | insula |

#### B.2 BrainNetCNN hyperparameter tuning and results

##### B.2.1 Model Hyperparameter Tuning

The BrainNetCNN models follow the layer order: E2E, E2N, N2G and a fully connected layer. Leaky ReLU activation and dropout were added after E2E, E2N and N2G layers. After taking out the test set, the rest of the data was split into training and validation sets with a ratio of 4:1 (this split is for hyperparameter tuning only and not used in the main analysis). Bayesian optimization was used to tune the hyperparameters within user-specified ranges of values based on the validation set. We tuned the number of filters for E2E (4 - 64 filters), E2N (4 - 128 filters), N2G (4 - 256 filters), dropout rate (0 – 0.95), Leaky ReLU scale (0 - 1), learning rate (1e-4 - 1e-2), L2 regularisation (1e-10 - 1e-4).

Table 2: Hyperparameters for the BrainNetCNN model. Under "Model Structure", the numbers represent the number of filters or node for each layer, with all the model following the same layer order: E2E, E2N, N2G and a fully connected layer

| Prediction Task | Model Structure | Dropout rate | Leaky ReLU scale | Learning rate | L2 regularisation | Optimizer |
|---|---|---|---|---|---|---|
| Sex | 22, 126, 240, 2 | 8.25 e-5 | 0.0089 | 0.0015 | 3.37e-9 | Adam |
| Age | 33, 125, 229, 1 | 0.0072 | 0.0573 | 0.0017 | 1.60e-10 | Adam |
| *g*-factor | 10, 95, 247, 1 | 1.43e-5 | 0.0383 | 0.0011 | 1.72e-6 | Adam |
| MHQ-factor | 10, 103, 186, 1 | 0.0021 | 3.38e-5 | 0.0010 | 1.18e-8 | Adam |

##### B.2.2 Results with hyperparameter tuning implemented

| **Weights** | MD | FA | SC | OD | ISOVF | ICVF |
|---|---|---|---|---|---|---|
| **Validation** | 79.07 (0.72) | 82.22 (0.63) | 89.14 (1.28) | 82.57 (1.41) | 82.34 (0.85) | 81.22 (0.68) |
| **Training** | 90.74 (3.27) | 93.14 (3.57) | 96.77 (2.93) | 93.88 (3.76) | 98.42 (3.16) | 94.31 (3.99) |
| **Test** | 77.90 (0.42) | 81.43 (0.64) | 86.81 (0.82) | 81.82 (1.17) | 82.86 (0.88) | 79.46 (1.29) |

(a) Sex prediction accuracies (mean percentage with standard deviation in brackets) with BrainNetCNN model for different connectivity weightings

| **Weights** | MD | FA | SC | OD | ISOVF | ICVF |
|---|---|---|---|---|---|---|
| **Validation** | 4.610 (0.662) | 4.646 (0.664) | 4.146 (0.731) | 4.345 (0.705) | 4.290 (0.717) | 4.644 (0.645) |
| **Training** | 3.495 (0.825) | 3.728 (0.806) | 3.523 (0.814) | 3.561 (0.820) | 3.445 (0.832) | 3.950 (0.768) |
| **Test** | 4.670 (0.645) | 4.725 (0.655) | 4.179 (0.711) | 4.533 (0.668) | 4.446 (0.684) | 4.617 (0.648) |

(b) Age prediction performance (mean absolute errors with correlations in brackets) with BrainNetCNN model for different connectivity weightings

| **Weights** | MD | FA | SC | OD | ISOVF | ICVF |
|---|---|---|---|---|---|---|
| **Validation** | 0.234 (0.757) | 0.235 (0.754) | 0.253 (0.752) | 0.236 (0.757) | 0.237 (0.759) | 0.237 (0.757) |
| **Training** | 0.271 (0.750) | 0.264 (0.750) | 0.312 (0.741) | 0.290 (0.744) | 0.270 (0.751) | 0.282 (0.747) |
| **Test** | 0.208 (0.767) | 0.223 (0.765) | 0.222 (0.769) | 0.213 (0.768) | 0.22 (0.766) | 0.218 (0.767) |

(c) *g*-factor prediction performance (correlations with mean absolute error in brackets) with BrainNetCNN model with age and sex covariates for different connectivity weightings

| **Weights** | MD | FA | SC | OD | ISOVF | ICVF |
|---|---|---|---|---|---|---|
| **Validation** | 0.224 (0.760) | 0.226 (0.764) | 0.227 (0.762) | 0.227 (0.762) | 0.225 (0.764) | 0.227 (0.767) |
| **Training** | 0.247 (0.756) | 0.244 (0.761) | 0.258 (0.755) | 0.241 (0.760) | 0.246 (0.760) | 0.254 (0.761) |
| **Test** | 0.248 (0.764) | 0.246 (0.769) | 0.240 (0.768) | 0.243 (0.768) | 0.239 (0.769) | 0.244 (0.772) |

(d) MHQ-factor prediction performance (correlations with mean absolute error in brackets) with BrainNetCNN model with age and sex covariates for different connectivity weightings

Table 3: Prediction performances with BrainNetCNN model of four different prediction tasks based on different connectivity weightings

#### B.3 Prediction performances of different classical machine learning models

##### B.3.1 Linear Ridge model

| **Weights** | MD | FA | SC | OD | ISOVF | ICVF |
|---|---|---|---|---|---|---|
| **Validation** | 80.91 (0.43) | 84.15 (0.88) | 89.67 (1.04) | 85.20 (0.63) | 84.12 (0.58) | 83.89 (0.45) |
| **Training** | 93.79 (5.03) | 94.43 (0.43) | 93.75 (1.40) | 92.94 (2.75) | 91.52 (2.09) | 97.80 (2.69) |
| **Test** | 80.91 (0.55) | 84.44 (0.32) | 87.54 (0.48) | 84.52 (0.40) | 84.68 (0.59) | 83.38 (0.46) |

(a) Sex prediction accuracies (mean percentage with standard deviation in brackets) with Linear Ridge model for different connectivity weightings

| **Weights** | MD | FA | SC | OD | ISOVF | ICVF |
|---|---|---|---|---|---|---|
| **Validation** | 4.472 (0.680) | 4.481 (0.679) | 4.221 (0.713) | 4.422 (0.688) | 4.204 (0.712) | 4.549 (0.668) |
| **Training** | 3.103 (0.855) | 2.968 (0.866) | 3.773 (0.779) | 3.668 (0.795) | 3.711 (0.789) | 3.085 (0.856) |
| **Test** | 4.465 (0.683) | 4.624 (0.661) | 4.258 (0.698) | 4.534 (0.665) | 4.250 (0.696) | 4.408 (0.693) |

(b) Age prediction performance (mean absolute errors with correlations in brackets) with Linear Ridge model for different connectivity weightings

| **Weights** | MD | FA | SC | OD | ISOVF | ICVF |
|---|---|---|---|---|---|---|
| **Validation** | 0.255 (0.753) | 0.256 (0.754) | 0.270 (0.748) | 0.267 (0.750) | 0.259 (0.751) | 0.256 (0.753) |
| **Training** | 0.318 (0.743) | 0.323 (0.744) | 0.330 (0.735) | 0.347 (0.734) | 0.330 (0.735) | 0.328 (0.742) |
| **Test** | 0.253 (0.761) | 0.262 (0.762) | 0.248 (0.764) | 0.254 (0.759) | 0.249 (0.759) | 0.258 (0.762) |

(c) $g$-factor prediction performance (correlations with mean absolute error in brackets) with Linear Ridge model with age and sex covariates for different connectivity weightings

| **Weights** | MD | FA | SC | OD | ISOVF | ICVF |
|---|---|---|---|---|---|---|
| **Validation** | 0.220 (0.770) | 0.216 (0.771) | 0.223 (0.767) | 0.225 (0.768) | 0.223 (0.769) | 0.220 (0.772) |
| **Training** | 0.320 (0.752) | 0.339 (0.749) | 0.281 (0.753) | 0.318 (0.749) | 0.299 (0.753) | 0.336 (0.75) |
| **Test** | 0.244 (0.777) | 0.240 (0.779) | 0.244 (0.772) | 0.247 (0.774) | 0.241 (0.775) | 0.242 (0.779) |

(d) MHQ-factor prediction performance (correlations with mean absolute error in brackets) with Linear Ridge model with age and sex covariates for different connectivity weightings

Table 4: Prediction performances with Linear Ridge model of four different prediction tasks based on different connectivity weightings

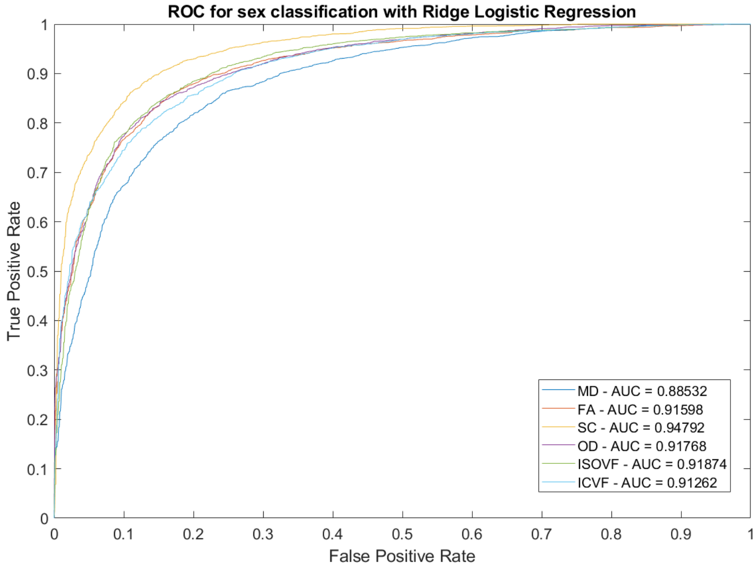


Figure 2: AUC and ROC curve for sex classification with Ridge Logistic Regression

###### B.3.2 Linear LASSO model

| Weights | MD | FA | SC | OD | ISOVF | ICVF |
|---|---|---|---|---|---|---|
| **Validation** | 79.97 (0.50) | 82.81 (0.94) | 88.22 (1.42) | 85.36 (0.60) | 84.24 (0.34) | 81.79 (0.63) |
| **Training** | 86.62 (1.01) | 90.30 (1.03) | 90.41 (0.29) | 92.07 (0.78) | 91.88 (1.28) | 90.26 (1.86) |
| **Test** | 79.80 (0.68) | 82.70 (0.57) | 85.90 (0.68) | 84.76 (0.53) | 84.65 (0.25) | 80.80 (0.97) |

(a) Sex prediction accuracies (mean percentage with standard deviation in brackets) with Linear LASSO model for different connectivity weightings

| Weights | MD | FA | SC | OD | ISOVF | ICVF |
|---|---|---|---|---|---|---|
| **Validation** | 4.459 (0.678) | 4.326 (0.693) | 4.238 (0.710) | 4.374 (0.709) | 4.224 (0.709) | 4.469 (0.667) |
| **Training** | 3.936 (0.774) | 3.652 (0.805) | 3.897 (0.763) | 3.550 (0.795) | 3.708 (0.788) | 3.766 (0.792) |
| **Test** | 4.456 (0.671) | 4.382 (0.680) | 4.282 (0.695) | 4.571 (0.680) | 4.274 (0.693) | 4.384 (0.681) |

(b) Age prediction performance (mean absolute errors with correlations in brackets) with Linear LASSO model for different connectivity weightings

| Weights | MD | FA | SC | OD | ISOVF | ICVF |
|---|---|---|---|---|---|---|
| **Validation** | 0.241 (0.756) | 0.223 (0.761) | 0.268 (0.749) | 0.256 (0.754) | 0.252 (0.752) | 0.237 (0.756) |
| **Training** | 0.339 (0.733) | 0.364 (0.725) | 0.335 (0.733) | 0.357 (0.728) | 0.312 (0.740) | 0.321 (0.737) |
| **Test** | 0.243 (0.760) | 0.241 (0.762) | 0.248 (0.765) | 0.244 (0.761) | 0.240 (0.761) | 0.242 (0.761) |

(c) $g$-factor prediction performance (correlations with mean absolute error in brackets) with Linear LASSO model with age and sex covariates for different connectivity weightings

| Weights | MD | FA | SC | OD | ISOVF | ICVF |
|---|---|---|---|---|---|---|
| **Validation** | 0.223 (0.769) | 0.211 (0.772) | 0.226 (0.768) | 0.214 (0.769) | 0.228 (0.768) | 0.219 (0.769) |
| **Training** | 0.267 (0.758) | 0.328 (0.739) | 0.279 (0.755) | 0.312 (0.744) | 0.275 (0.756) | 0.308 (0.746) |
| **Test** | 0.240 (0.775) | 0.224 (0.781) | 0.237 (0.776) | 0.230 (0.779) | 0.247 (0.773) | 0.235 (0.777) |

(d) MHQ-factor prediction performance (correlations with mean absolute error in brackets) with Linear Ridge model with age and sex covariates for different connectivity weightings

Table 5: Prediction performances with Linear LASSO model of four different prediction tasks based on different connectivity weightings

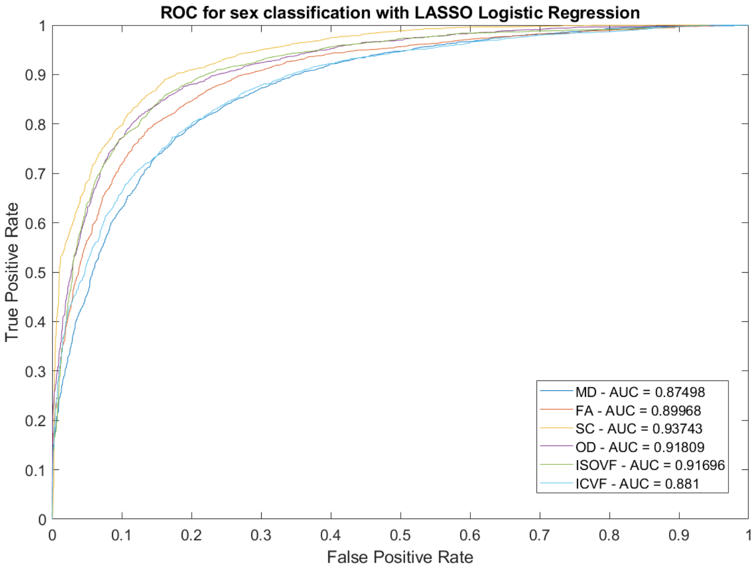


Figure 3: AUC and ROC curve for sex classification with LASSO Logistic Regression

##### B.3.3 Linear SVM model

| **Weights** | MD | FA | SC | OD | ISOVF | ICVF |
|---|---|---|---|---|---|---|
| **Validation** | 82.05 (0.80) | 84.63 (0.82) | 89.77 (0.91) | 85.83 (0.43) | 84.37 (0.65) | 85.55 (1.13) |
| **Training** | 96.05 (1.27) | 94.06 (2.46) | 93.90 (0.89) | 92.06 (1.37) | 92.15 (1.50) | 98.33 (0.51) |
| **Test** | 80.65 (0.62) | 83.56 (1.12) | 87.27 (0.56) | 84.68 (0.35) | 84.18 (0.52) | 84.37 (0.82) |

(a) Sex prediction accuracies (mean percentage with standard deviation in brackets) with Linear SVM model for different connectivity weightings

| **Weights** | MD | FA | SC | OD | ISOVF | ICVF |
|---|---|---|---|---|---|---|
| **Validation** | 4.574 (0.660) | 4.450 (0.680) | 4.318 (0.698) | 4.428 (0.683) | 4.369 (0.690) | 4.614 (0.654) |
| **Training** | 3.214 (0.806) | 3.009 (0.827) | 3.975 (0.738) | 3.156 (0.816) | 3.147 (0.817) | 3.167 (0.81) |
| **Test** | 4.537 (0.664) | 4.649 (0.656) | 4.357 (0.684) | 4.527 (0.662) | 4.453 (0.679) | 4.500 (0.672) |

(b) Age prediction performance (mean absolute errors with correlations in brackets) with Linear SVM model for different connectivity weightings

| **Weights** | MD | FA | SC | OD | ISOVF | ICVF |
|---|---|---|---|---|---|---|
| **Validation** | 0.257 (0.752) | 0.256 (0.752) | 0.270 (0.745) | 0.268 (0.748) | 0.260 (0.748) | 0.257 (0.752) |
| **Training** | 0.311 (0.738) | 0.313 (0.740) | 0.316 (0.730) | 0.330 (0.729) | 0.322 (0.729) | 0.319 (0.737) |
| **Test** | 0.257 (0.761) | 0.264 (0.762) | 0.246 (0.763) | 0.247 (0.760) 0.250 (0.759) | 0.261 (0.762) | |

(c) *g*-factor prediction performance (correlations with mean absolute error in brackets) with Linear SVM model with age and sex covariates for different connectivity weightings

| **Weights** | MD | FA | SC | OD | ISOVF | ICVF |
|---|---|---|---|---|---|---|
| **Validation** | 0.218 (0.743) | 0.215 (0.744) | 0.224 (0.794) | 0.226 (0.740) | 0.221 (0.743) | 0.216 (0.744) |
| **Training** | 0.299 (0.717) | 0.304 (0.717) | 0.286 (0.750) | 0.309 (0.711) | 0.272 (0.724) | 0.306 (0.716) |
| **Test** | 0.245 (0.745) | 0.245 (0.746) | 0.230 (0.794) | 0.248 (0.743) | 0.240 (0.745) | 0.244 (0.747) |

(d) MHQ-factor prediction performance (correlations with mean absolute error in brackets) with Linear SVM model with age and sex covariates for different connectivity weightings

Table 6: Prediction performances with Linear SVM model of four different prediction tasks based on different connectivity weightings

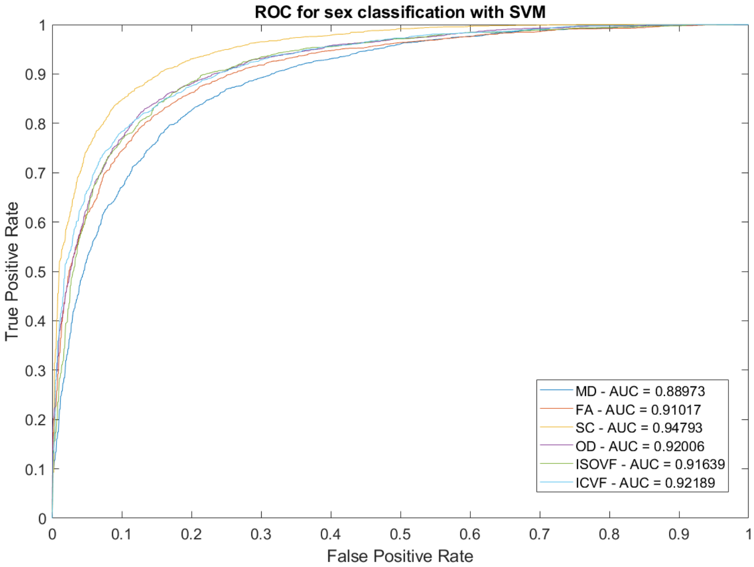


Figure 4: AUC and ROC curve for sex classification with SVM

##### B.3.4 Kernel Ridge model

| Weights | MD | FA | SC | OD | ISOVF | ICVF |
|---|---|---|---|---|---|---|
| **Validation** | 79.62 (0.45) | 81.62 (0.85) | 86.44 (1.30) | 83.30 (0.71) | 82.53 (0.65) | 80.50 (0.96) |
| **Training** | 90.46 (0.12) | 91.16 (0.23) | 88.09 (0.36) | 91.56 (0.80) | 89.94 (0.42) | 91.29 (0.59) |
| **Test** | 79.50 (0.66) | 80.86 (0.22) | 84.28 (0.36) | 82.89 (0.46) | 82.73 (0.43) | 79.84 (0.76) |

(a) Sex prediction accuracies (mean percentage with standard deviation in brackets) with Kernel Ridge model for different connectivity weightings

| Weights | MD | FA | SC | OD | ISOVF | ICVF |
|---|---|---|---|---|---|---|
| **Validation** | 4.550 (0.653) | 4.550 (0.651) | 4.402 (0.685) | 4.313 (0.693) | 4.244 (0.707) | 4.655 (0.632) |
| **Training** | 3.856 (0.766) | 3.902 (0.761) | 4.271 (0.707) | 3.761 (0.778) | 3.768 (0.778) | 3.990 (0.746) |
| **Test** | 4.485 (0.655) | 4.491 (0.660) | 4.434 (0.669) | 4.378 (0.671) | 4.255 (0.693) | 4.563 (0.642) |

(b) Age prediction performance (mean absolute errors with correlations in brackets) with Kernel Ridge model for different connectivity weightings

| Weights | MD | FA | SC | OD | ISOVF | ICVF |
|---|---|---|---|---|---|---|
| **Validation** | 0.251 (0.753) | 0.234 (0.769) | 0.245 (0.759) | 0.188 (0.804) | 0.178 (0.801) | 0.241 (0.767) |
| **Training** | 0.307 (0.741) | 0.337 (0.731) | 0.354 (0.728) | 0.463 (0.690) | 0.450 (0.699) | 0.344 (0.728) |
| **Test** | 0.252 (0.759) | 0.241 (0.773) | 0.257 (0.768) | 0.191 (0.804) | 0.176 (0.806) | 0.241 (0.773) |

(c) $g$-factor prediction performance (correlations with mean absolute error in brackets) with Kernel Ridge model with age and sex covariates for different connectivity weightings

| Weights | MD | FA | SC | OD | ISOVF | ICVF |
|---|---|---|---|---|---|---|
| **Validation** | 0.220 (0.769) | 0.208 (0.785) | 0.207 (0.774) | 0.188 (0.789) | 0.218 (0.774) | 0.220 (0.769) |
| **Training** | 0.274 (0.757) | 0.301 (0.743) | 0.269 (0.758) | 0.352 (0.725) | 0.303 (0.744) | 0.284 (0.754) |
| **Test** | 0.245 (0.775) | 0.226 (0.793) | 0.229 (0.780) | 0.207 (0.797) | 0.218 (0.790) | 0.245 (0.774) |

(d) MHQ-factor prediction performance (correlations with mean absolute error in brackets) with Kernel Ridge model with age and sex covariates for different connectivity weightings

Table 7: Prediction performances with Kernel Ridge model of four different prediction tasks based on different connectivity weightings

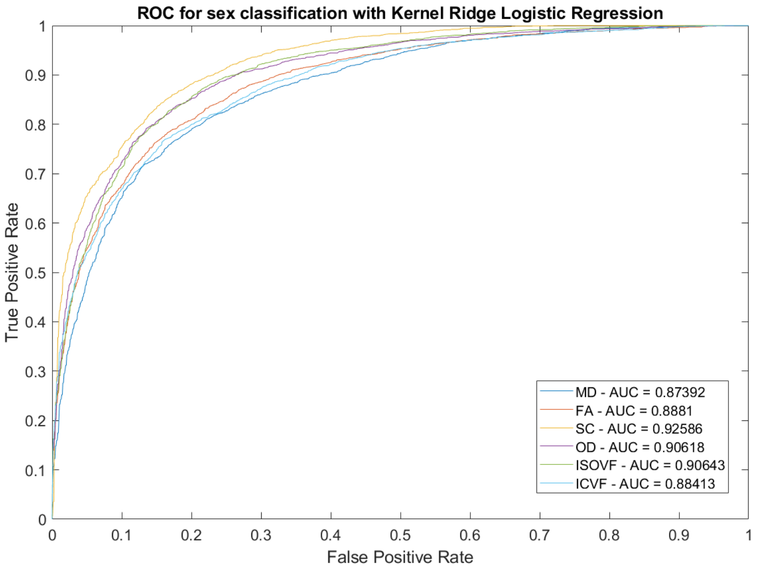


Figure 5: AUC and ROC curve for sex classification with Kernel Ridge Logistic Regression

##### B.4 Consistency for the gradient attribution maps and beta coefficients

###### B.4.1 Consistency across folds

Table 8: The consistency for the gradient attribution maps across folds for BrainNetCNN and the consistency for the beta coefficients across folds for linear ridge regressions of different prediction tasks based on streamline count, illustrated using Spearman's correlation

| | Streamline Count Structural Connectivity | | | | | | | | | |
|---|---|---|---|---|---|---|---|---|---|---|
| | BrainNetCNN | | | | | Linear Ridge Regression | | | | |
| Sex Classification | | CV1 | CV2 | CV3 | CV4 | | CV1 | CV2 | CV3 | CV4 |
| | CV2 | 0.803 | 0.799 | 0.798 | 0.802 | CV2 | 0.840 | 0.827 | 0.861 | 0.782 |
| | CV3 | | 0.801 | 0.792 | 0.803 | CV3 | | 0.859 | 0.857 | 0.719 |
| | CV4 | | | 0.815 | 0.799 | CV4 | | | 0.867 | 0.705 |
| | CV5 | | | | 0.812 | CV5 | | | | 0.736 |
| Age Prediction | | CV1 | CV2 | CV3 | CV4 | | CV1 | CV2 | CV3 | CV4 |
| | CV2 | 0.748 | 0.760 | 0.754 | 0.762 | CV2 | 0.816 | 0.810 | 0.823 | 0.827 |
| | CV3 | | 0.762 | 0.760 | 0.753 | CV3 | | 0.827 | 0.830 | 0.830 |
| | CV4 | | | 0.776 | 0.760 | CV4 | | | 0.832 | 0.826 |
| | CV5 | | | | 0.773 | CV5 | | | | 0.841 |
| g-factor Prediction | | CV1 | CV2 | CV3 | CV4 | | CV1 | CV2 | CV3 | CV4 |
| | CV2 | 0.586 | 0.580 | 0.559 | 0.581 | CV2 | 0.721 | 0.703 | 0.696 | 0.714 |
| | CV3 | | 0.579 | 0.581 | 0.603 | CV3 | | 0.706 | 0.673 | 0.709 |
| | CV4 | | | 0.563 | 0.563 | CV4 | | | 0.672 | 0.692 |
| | CV5 | | | | 0.559 | CV5 | | | | 0.667 |
| MHQ-factor Prediction | | CV1 | CV2 | CV3 | CV4 | | CV1 | CV2 | CV3 | CV4 |
| | CV2 | 0.528 | 0.516 | 0.510 | 0.485 | CV2 | 0.676 | 0.665 | 0.681 | 0.652 |
| | CV3 | | 0.555 | 0.560 | 0.530 | CV3 | | 0.691 | 0.710 | 0.689 |
| | CV4 | | | 0.537 | 0.497 | CV4 | | | 0.693 | 0.657 |
| | CV5 | | | | 0.502 | CV5 | | | | 0.692 |

Table 9: The consistency for the gradient attribution maps across folds for BrainNetCNN and the consistency for the beta coefficients across folds for linear ridge regressions of different prediction tasks based on fractional anisotropy, illustrated using Spearman's correlation

| | Fractional Anisotropy Structural Connectivity | | | | | | | | | |
|---|---|---|---|---|---|---|---|---|---|---|
| | BrainNetCNN | | | | | Linear Ridge Regression | | | | |
| Sex Classification | | CV1 | CV2 | CV3 | CV4 | | CV1 | CV2 | CV3 | CV4 |
| | CV2 | 0.831 | 0.821 | 0.811 | 0.823 | CV2 | 0.872 | 0.863 | 0.867 | 0.873 |
| | CV3 | | 0.819 | 0.831 | 0.833 | CV3 | | 0.854 | 0.862 | 0.872 |
| | CV4 | | | 0.820 | 0.824 | CV4 | | | 0.859 | 0.863 |
| | CV5 | | | | 0.818 | CV5 | | | | 0.863 |
| Age Prediction | | CV1 | CV2 | CV3 | CV4 | | CV1 | CV2 | CV3 | CV4 |
| | CV2 | 0.832 | 0.818 | 0.818 | 0.833 | CV2 | 0.716 | 0.709 | 0.731 | 0.737 |
| | CV3 | | 0.807 | 0.813 | 0.834 | CV3 | | 0.718 | 0.714 | 0.734 |
| | CV4 | | | 0.808 | 0.819 | CV4 | | | 0.705 | 0.719 |
| | CV5 | | | | 0.807 | CV5 | | | | 0.740 |
| g-factor Prediction | | CV1 | CV2 | CV3 | CV4 | | CV1 | CV2 | CV3 | CV4 |
| | CV2 | 0.648 | 0.605 | 0.603 | 0.622 | CV2 | 0.763 | 0.749 | 0.720 | 0.742 |
| | CV3 | | 0.630 | 0.616 | 0.638 | CV3 | | 0.760 | 0.759 | 0.786 |
| | CV4 | | | 0.601 | 0.624 | CV4 | | | 0.752 | 0.751 |
| | CV5 | | | | 0.617 | CV5 | | | | 0.747 |
| MHQ-factor Prediction | | CV1 | CV2 | CV3 | CV4 | | CV1 | CV2 | CV3 | CV4 |
| | CV2 | 0.592 | 0.571 | 0.598 | 0.558 | CV2 | 0.684 | 0.656 | 0.711 | 0.664 |
| | CV3 | | 0.593 | 0.638 | 0.598 | CV3 | | 0.691 | 0.708 | 0.698 |
| | CV4 | | | 0.608 | 0.566 | CV4 | | | 0.708 | 0.685 |
| | CV5 | | | | 0.595 | CV5 | | | | 0.699 |

##### B.4.2 Consistency across methods

Table 10: The consistency between the gradient attribution maps from BrainNetCNN and the beta coefficients from classical methods of different prediction tasks based on streamline count and fractional anisotropy, illustrated using Spearman's correlation

| Connectivity Weight | Streamline Count | | | | Fractional Anisotropy | | | |
|---|---|---|---|---|---|---|---|---|
| Prediction Type | Method | Ridge Regression | LASSO Regression | SVM | Method | Ridge Regression | LASSO Regression | SVM |
| Sex Classification | CV1 | 0.736 | 0.660 | 0.696 | CV1 | 0.852 | 0.868 | 0.781 |
| | CV2 | 0.770 | 0.699 | 0.736 | CV2 | 0.848 | 0.865 | 0.812 |
| | CV3 | 0.750 | 0.653 | 0.723 | CV3 | 0.834 | 0.830 | 0.774 |
| | CV4 | 0.759 | 0.677 | 0.729 | CV4 | 0.848 | 0.862 | 0.837 |
| | CV5 | 0.637 | 0.607 | 0.613 | CV5 | 0.826 | 0.833 | 0.809 |
| Age Prediction | CV1 | 0.708 | 0.741 | 0.668 | CV1 | 0.509 | 0.825 | 0.630 |
| | CV2 | 0.710 | 0.745 | 0.682 | CV2 | 0.522 | 0.769 | 0.623 |
| | CV3 | 0.682 | 0.735 | 0.647 | CV3 | 0.490 | 0.847 | 0.618 |
| | CV4 | 0.699 | 0.741 | 0.671 | CV4 | 0.521 | 0.847 | 0.632 |
| | CV5 | 0.713 | 0.745 | 0.670 | CV5 | 0.511 | 0.830 | 0.627 |
| g-factor Prediction | CV1 | 0.676 | 0.455 | 0.535 | CV1 | 0.775 | 0.786 | 0.596 |
| | CV2 | 0.695 | 0.531 | 0.559 | CV2 | 0.781 | 0.798 | 0.620 |
| | CV3 | 0.679 | 0.491 | 0.534 | CV3 | 0.776 | 0.588 | 0.606 |
| | CV4 | 0.674 | 0.515 | 0.533 | CV4 | 0.788 | 0.771 | 0.631 |
| | CV5 | 0.682 | 0.468 | 0.545 | CV5 | 0.780 | 0.805 | 0.609 |
| MHQ-factor Prediction | CV1 | 0.693 | 0.502 | 0.496 | CV1 | 0.763 | 0.787 | 0.574 |
| | CV2 | 0.677 | 0.527 | 0.545 | CV2 | 0.773 | 0.792 | 0.569 |
| | CV3 | 0.671 | 0.521 | 0.523 | CV3 | 0.769 | 0.809 | 0.563 |
| | CV4 | 0.661 | 0.572 | 0.486 | CV4 | 0.771 | 0.794 | 0.571 |
| | CV5 | 0.648 | 0.523 | 0.511 | CV5 | 0.772 | 0.801 | 0.568 |

##### B.4.3 Top regions for different prediction tasks of BrainNetCNN and Ridge Regression

Table 11: Top 20 predictive regions based on streamline count and fractional anisotropy connectivities for four different prediction tasks, selected by BrainNetCNN models and linear ridge regression models. Overlapping regions are highlighted in bold text

| Connectivity Weight | Streamline Count | | Fractional Anisotropy | |
|---|---|---|---|---|
| Top Regions | BrainNetCNN | Ridge Regression | BrainNetCNN | Ridge Regression |
| Sex Classification | **ctx-lh-precuneus** | **Right-Putamen** | **Right-Thalamus-Proper** | **Right-Thalamus-Proper** |
| | **ctx-rh-superiortemporal** | **Left-Putamen** | **Right-Caudate** | **Right-Putamen** |
| | **ctx-rh-precuneus** | ctx-rh-superiorfrontal | **Right-Putamen** | ctx-lh-precuneus |
| | **ctx-rh-insula** | **Left-Thalamus-Proper** | **ctx-lh-isthmuscingulate** | **ctx-lh-isthmuscingulate** |
| | **Right-Putamen** | **Right-Thalamus-Proper** | **ctx-lh-superiorfrontal** | **Left-Putamen** |
| | **Right-Thalamus-Proper** | **ctx-lh-superiorfrontal** | ctx-lh-rostralmiddlefrontal | **ctx-rh-superiorparietal** |
| | **ctx-rh-inferiortemporal** | **ctx-rh-precuneus** | **ctx-rh-insula** | **Right-Caudate** |
| | ctx-lh-supramarginal | **ctx-lh-precuneus** | ctx-rh-posteriorcingulate | **Right-VentralDC** |
| | ctx-rh-paracentral | ctx-rh-superiorparietal | **ctx-rh-inferiortemporal** | **ctx-rh-inferiortemporal** |
| | Right-Hippocampus | **ctx-rh-insula** | **Left-Putamen** | ctx-rh-postcentral |
| | **ctx-rh-supramarginal** | Left-Caudate | Left-Hippocampus | **Left-Thalamus-Proper** |
| | **Right-VentralDC** | ctx-lh-inferiortemporal | ctx-rh-isthmuscingulate | **ctx-rh-insula** |
| | **Left-Putamen** | **ctx-lh-isthmuscingulate** | **Right-VentralDC** | **ctx-lh-superiorfrontal** |
| | **Left-Thalamus-Proper** | **ctx-rh-inferiortemporal** | **ctx-rh-superiorparietal** | **Left-Caudate** |
| | **ctx-lh-superiortemporal** | **ctx-rh-superiortemporal** | **ctx-rh-precuneus** | **ctx-rh-precuneus** |
| | ctx-lh-medialorbitofrontal | **ctx-rh-supramarginal** | **Left-Caudate** | ctx-rh-precentral |
| | **ctx-lh-superiorfrontal** | ctx-lh-superiorparietal | **ctx-rh-superiorfrontal** | ctx-lh-superiorparietal |
| | **ctx-lh-isthmuscingulate** | **ctx-lh-superiortemporal** | ctx-rh-rostralmiddlefrontal | **ctx-rh-superiorfrontal** |
| | Right-Accumbens-area | ctx-rh-middletemporal | ctx-rh-lateralorbitofrontal | ctx-lh-inferiorparietal |
| | ctx-rh-temporalpole | **Right-VentralDC** | **Left-Thalamus-Proper** | ctx-rh-lateraloccipital |

Table 11: Top 20 predictive regions based on streamline count and fractional anisotropy connectivities for four different prediction tasks, selected by BrainNetCNN models and linear ridge regression models. Overlapping regions are highlighted in bold text (cont.)

| Connectivity Weight | Streamline Count | | Fractional Anisotropy | |
|---|---|---|---|---|
| Top Regions | BrainNetCNN | Ridge Regression | BrainNetCNN | Ridge Regression |
| Age Prediction | **Right-Thalamus-Proper** | **Right-Thalamus-Proper** | **Left-Thalamus-Proper** | **Right-Putamen** |
| | Right-Accumbens-area | **Left-Putamen** | **Right-Putamen** | **Left-Thalamus-Proper** |
| | **Right-VentralDC** | **ctx-rh-precuneus** | **Right-Thalamus-Proper** | **ctx-rh-superiorfrontal** |
| | ctx-rh-medialorbitofrontal | **ctx-rh-superiorparietal** | Right-Caudate | **Right-Thalamus-Proper** |
| | **ctx-rh-precuneus** | **Right-Putamen** | Left-VentralDC | **Left-Putamen** |
| | **Left-Caudate** | ctx-rh-superiorfrontal | **Left-Putamen** | ctx-rh-superiorparietal |
| | **Right-Pallidum** | **Left-Caudate** | **ctx-lh-isthmuscingulate** | **ctx-lh-precuneus** |
| | **Left-Putamen** | **Right-VentralDC** | **ctx-rh-precuneus** | **ctx-rh-precuneus** |
| | Left-Hippocampus | Left-Thalamus-Proper | Left-Hippocampus | **Left-Caudate** |
| | **Left-VentralDC** | **ctx-lh-superiorparietal** | **Right-VentralDC** | **ctx-rh-insula** |
| | **ctx-rh-insula** | **Right-Pallidum** | **ctx-rh-superiorfrontal** | **ctx-lh-superiorfrontal** |
| | **ctx-rh-superiorparietal** | **Right-Caudate** | **ctx-rh-insula** | **ctx-lh-isthmuscingulate** |
| | ctx-lh-medialorbitofrontal | ctx-lh-precuneus | ctx-rh-isthmuscingulate | **Right-VentralDC** |
| | **Right-Caudate** | **Left-VentralDC** | **ctx-lh-superiorfrontal** | ctx-lh-superiortemporal |
| | **ctx-lh-superiortemporal** | **ctx-lh-superiortemporal** | **Left-Caudate** | ctx-rh-rostralmiddlefrontal |
| | Left-Pallidum | **ctx-rh-insula** | **ctx-lh-precuneus** | ctx-rh-middletemporal |
| | ctx-lh-middletemporal | ctx-lh-superiorfrontal | ctx-lh-insula | ctx-lh-middletemporal |
| | **ctx-lh-superiorparietal** | ctx-rh-inferiorparietal | Left-Pallidum | ctx-rh-inferiorparietal |
| | ctx-rh-isthmuscingulate | ctx-lh-isthmuscingulate | ctx-rh-parsopercularis | ctx-rh-precentral |
| | **Right-Putamen** | ctx-rh-superiortemporal | Right-Pallidum | ctx-rh-inferiortemporal |
| *g*-factor Prediction | **Right-Thalamus-Proper** | **Left-Putamen** | **ctx-rh-isthmuscingulate** | **Left-Thalamus-Proper** |
| | **Left-Putamen** | **Left-Thalamus-Proper** | **Right-VentralDC** | **Right-VentralDC** |
| | **Left-Hippocampus** | **Right-Putamen** | **ctx-rh-insula** | **ctx-rh-isthmuscingulate** |
| | **ctx-rh-superiorparietal** | **ctx-rh-superiorfrontal** | **Left-VentralDC** | ctx-lh-fusiform |
| | **ctx-rh-precuneus** | **Right-Thalamus-Proper** | **ctx-rh-inferiortemporal** | ctx-rh-caudalanteriorcingulate |
| | **ctx-lh-superiortemporal** | ctx-lh-superiorfrontal | Right-Caudate | ctx-lh-caudalanteriorcingulate |
| | **Left-Caudate** | **ctx-lh-superiorparietal** | ctx-lh-postcentral | **ctx-rh-lateraloccipital** |
| | ctx-lh-paracentral | **ctx-rh-superiorparietal** | **Right-Pallidum** | **Left-Pallidum** |
| | **ctx-lh-rostralmiddlefrontal** | **Left-Caudate** | **ctx-rh-lateraloccipital** | **ctx-rh-inferiortemporal** |
| | **ctx-lh-superiorparietal** | **ctx-lh-precuneus** | **Left-Pallidum** | **ctx-rh-insula** |
| | **ctx-lh-inferiorparietal** | **ctx-lh-inferiorparietal** | Left-Putamen | **Right-Pallidum** |
| | ctx-rh-parstriangularis | **Right-Caudate** | **ctx-rh-inferiorparietal** | **ctx-rh-fusiform** |
| | ctx-rh-isthmuscingulate | **ctx-rh-precuneus** | **ctx-rh-fusiform** | ctx-rh-posteriorcingulate |
| | **Right-Caudate** | **Left-Hippocampus** | ctx-rh-bankssts | **ctx-lh-isthmuscingulate** |
| | **ctx-lh-precuneus** | **ctx-rh-rostralmiddlefrontal** | ctx-rh-paracentral | **Left-VentralDC** |
| | **ctx-rh-superiorfrontal** | ctx-rh-inferiorparietal | **Left-Thalamus-Proper** | ctx-lh-precuneus |
| | ctx-rh-parsopercularis | **ctx-lh-rostralmiddlefrontal** | **ctx-lh-isthmuscingulate** | **ctx-rh-superiortemporal** |
| | **Left-Thalamus-Proper** | ctx-rh-lateraloccipital | **ctx-rh-superiortemporal** | ctx-rh-middletemporal |
| | **ctx-rh-rostralmiddlefrontal** | ctx-lh-precentral | ctx-lh-inferiorparietal | **ctx-rh-inferiorparietal** |
| | **Right-Putamen** | **ctx-lh-superiortemporal** | **Left-Hippocampus** | **Left-Hippocampus** |
| MHQ-factor Prediction | **ctx-lh-precuneus** | **Left-Putamen** | **Left-VentralDC** | **ctx-lh-posteriorcingulate** |
| | **Left-Putamen** | **Left-Thalamus-Proper** | **ctx-lh-posteriorcingulate** | **ctx-rh-isthmuscingulate** |
| | **ctx-rh-superiorparietal** | **Right-Putamen** | **ctx-rh-insula** | **Right-Pallidum** |
| | **ctx-rh-precuneus** | **ctx-rh-superiorparietal** | **ctx-lh-insula** | **Left-Pallidum** |
| | **ctx-lh-superiortemporal** | **Right-Thalamus-Proper** | **ctx-lh-middletemporal** | **Left-VentralDC** |
| | ctx-rh-superiortemporal | **Left-Caudate** | **ctx-rh-inferiortemporal** | ctx-rh-caudalanteriorcingulate |
| | ctx-lh-lingual | **ctx-lh-precuneus** | **ctx-rh-isthmuscingulate** | **ctx-rh-inferiortemporal** |
| | **Left-Caudate** | **ctx-rh-precuneus** | **ctx-lh-inferiorparietal** | **ctx-rh-insula** |
| | **ctx-lh-isthmuscingulate** | ctx-lh-superiorfrontal | **Right-Pallidum** | Right-Hippocampus |
| | **Right-Putamen** | **ctx-lh-superiorparietal** | **Left-Pallidum** | **ctx-rh-paracentral** |
| | **ctx-lh-fusiform** | **ctx-lh-isthmuscingulate** | ctx-rh-inferiorparietal | **ctx-rh-middletemporal** |
| | **ctx-lh-inferiortemporal** | ctx-rh-inferiorparietal | **ctx-rh-middletemporal** | **Right-VentralDC** |
| | Left-Hippocampus | **ctx-lh-superiortemporal** | **Right-VentralDC** | ctx-lh-lingual |
| | ctx-rh-lateralorbitofrontal | ctx-rh-superiorfrontal | ctx-rh-precuneus | **ctx-lh-inferiorparietal** |
| | ctx-lh-medialorbitofrontal | **ctx-lh-fusiform** | **ctx-lh-lateraloccipital** | **ctx-lh-lateraloccipital** |
| | **Right-Thalamus-Proper** | **ctx-lh-inferiortemporal** | **ctx-rh-lateraloccipital** | **ctx-lh-insula** |
| | **ctx-lh-superiorparietal** | ctx-lh-lateraloccipital | **ctx-rh-paracentral** | **ctx-lh-middletemporal** |
| | **Right-Caudate** | **Right-Caudate** | ctx-lh-parstriangularis | **ctx-lh-fusiform** |
| | **Left-Thalamus-Proper** | ctx-lh-inferiorparietal | **ctx-lh-fusiform** | ctx-rh-frontalpole |
| | ctx-lh-paracentral | ctx-lh-supramarginal | ctx-lh-medialorbitofrontal | **ctx-rh-lateraloccipital** |

##### B.4.4 Training Progress of BrainNetCNN models for different prediction tasks based on six different dMRI measures

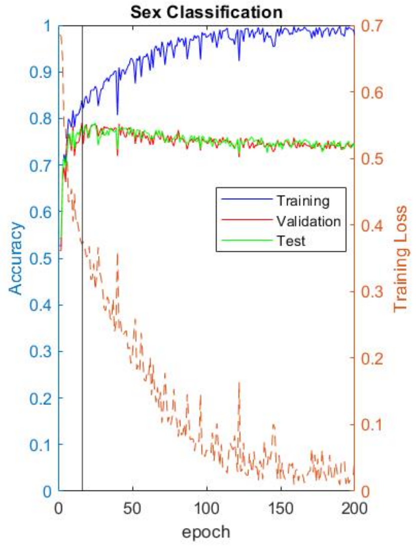


(a) MD

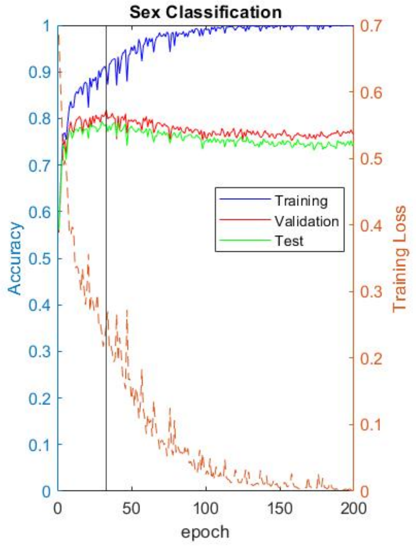


(b) FA

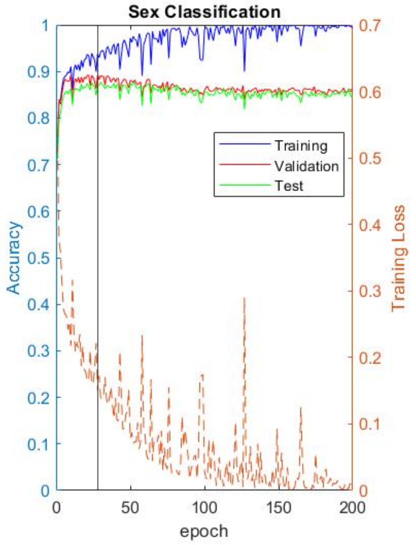


(c) SC

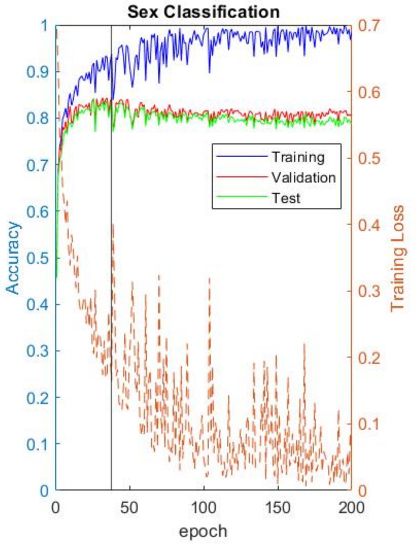


(d) OD

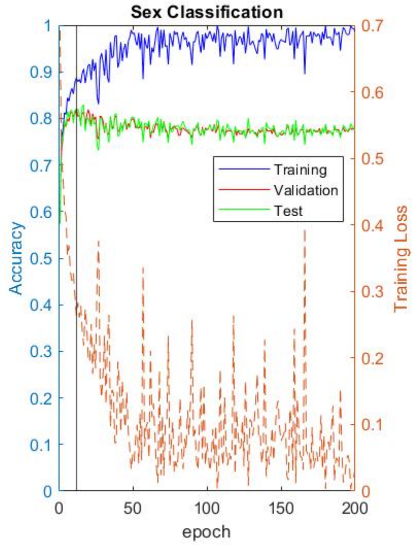


(e) ISOVF

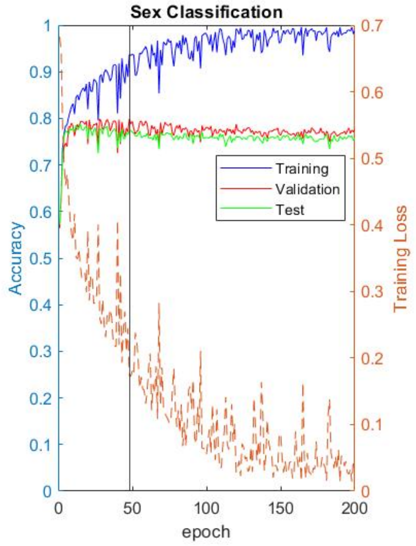


(f) ICVF

Figure 6: The BrainNetCNN models' training progress for sex classification for the fourth fold based on the six different dMRI measures. Training loss is the cross-entropy loss

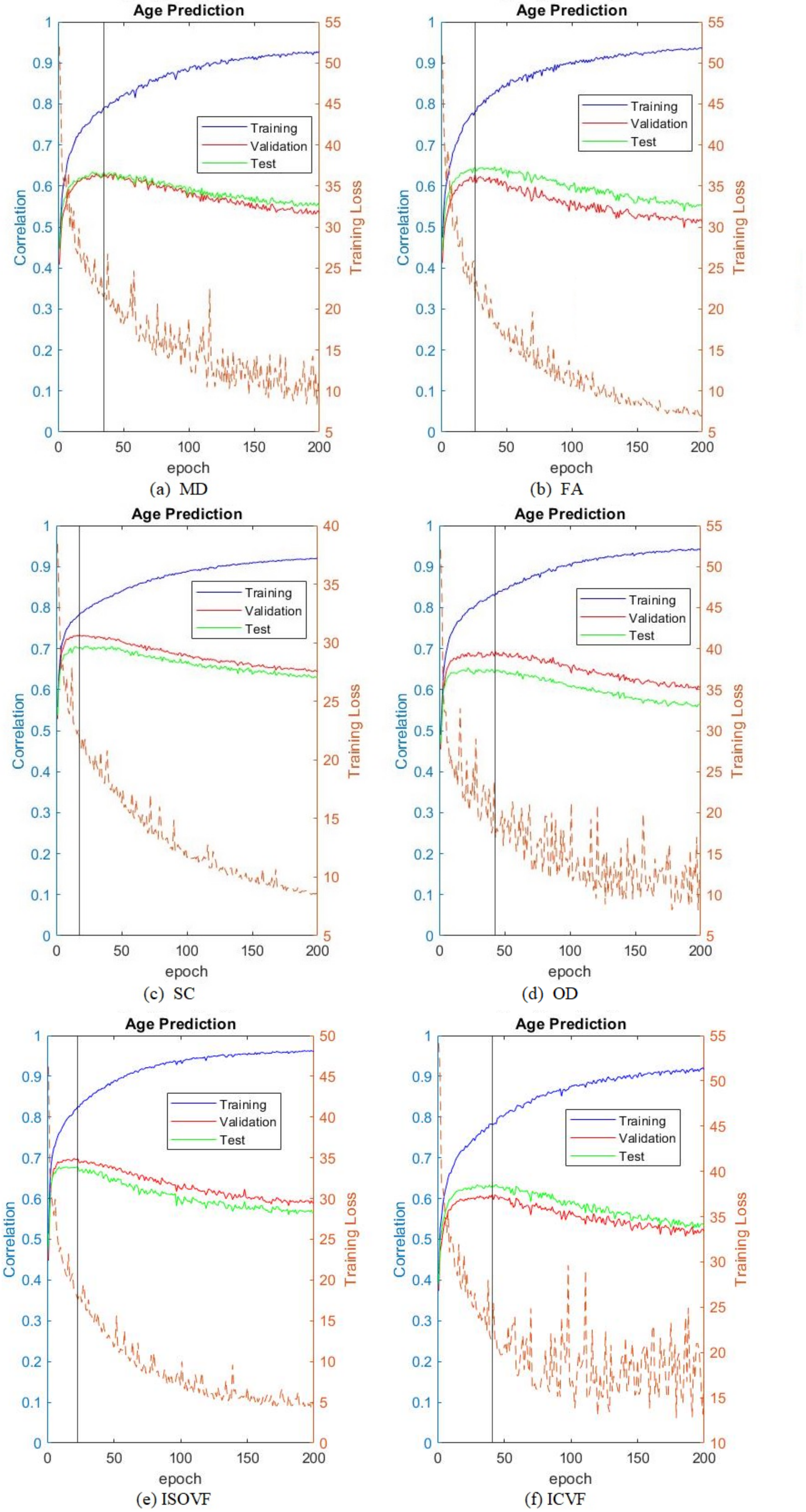


(a) MD (b) FA (c) SC (d) OD (e) ISOVF (f) ICVF

Figure 7: The BrainNetCNN models' training progress for age prediction for the fourth fold based on the six different dMRI measures. Training loss is the mean square loss

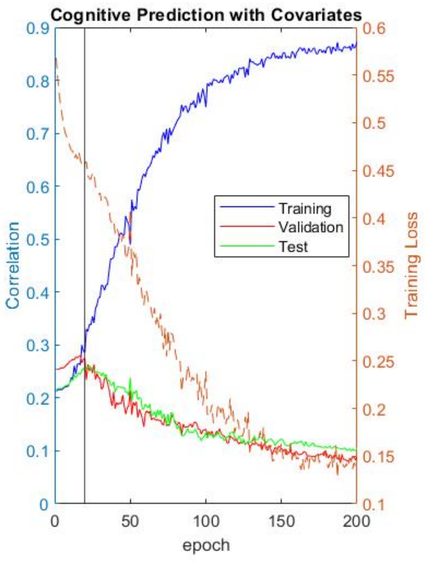

(a) MD

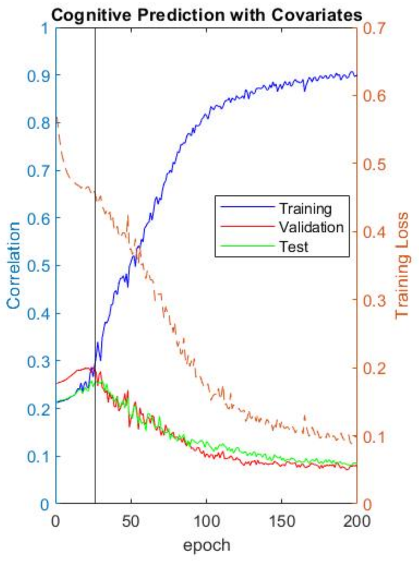

(b) FA

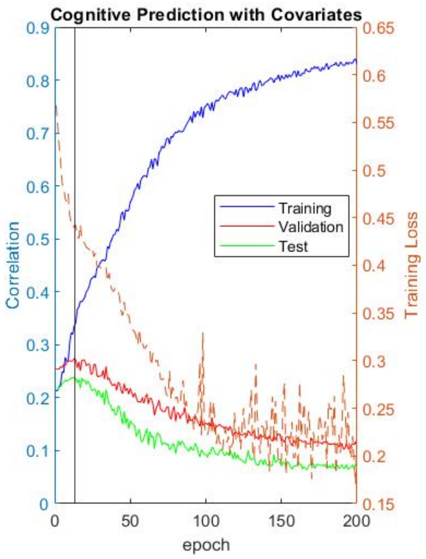

(c) SC

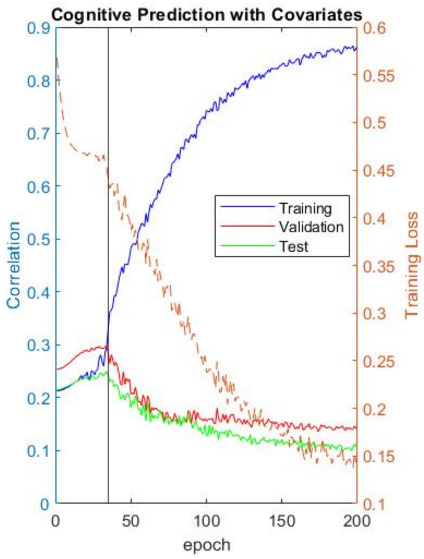

(d) OD

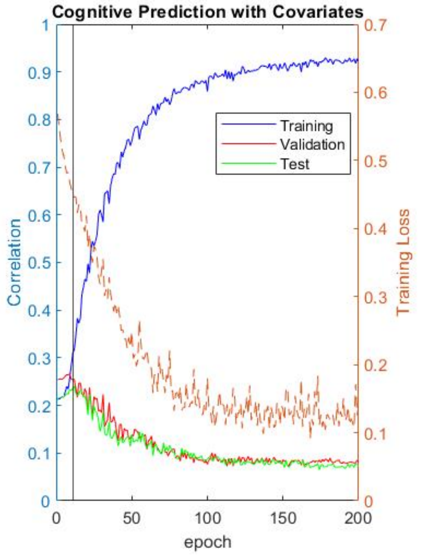

(e) ISOVF

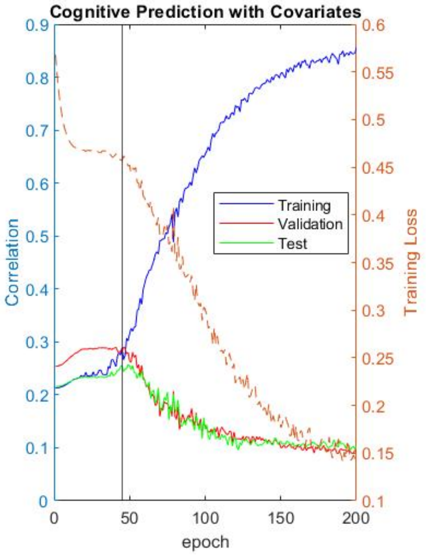

(f) ICVF

Figure 8: The BrainNetCNN models' training progress for $g$-factor prediction for the fourth fold based on the six different dMRI measures. Training loss is the cross-entropy loss

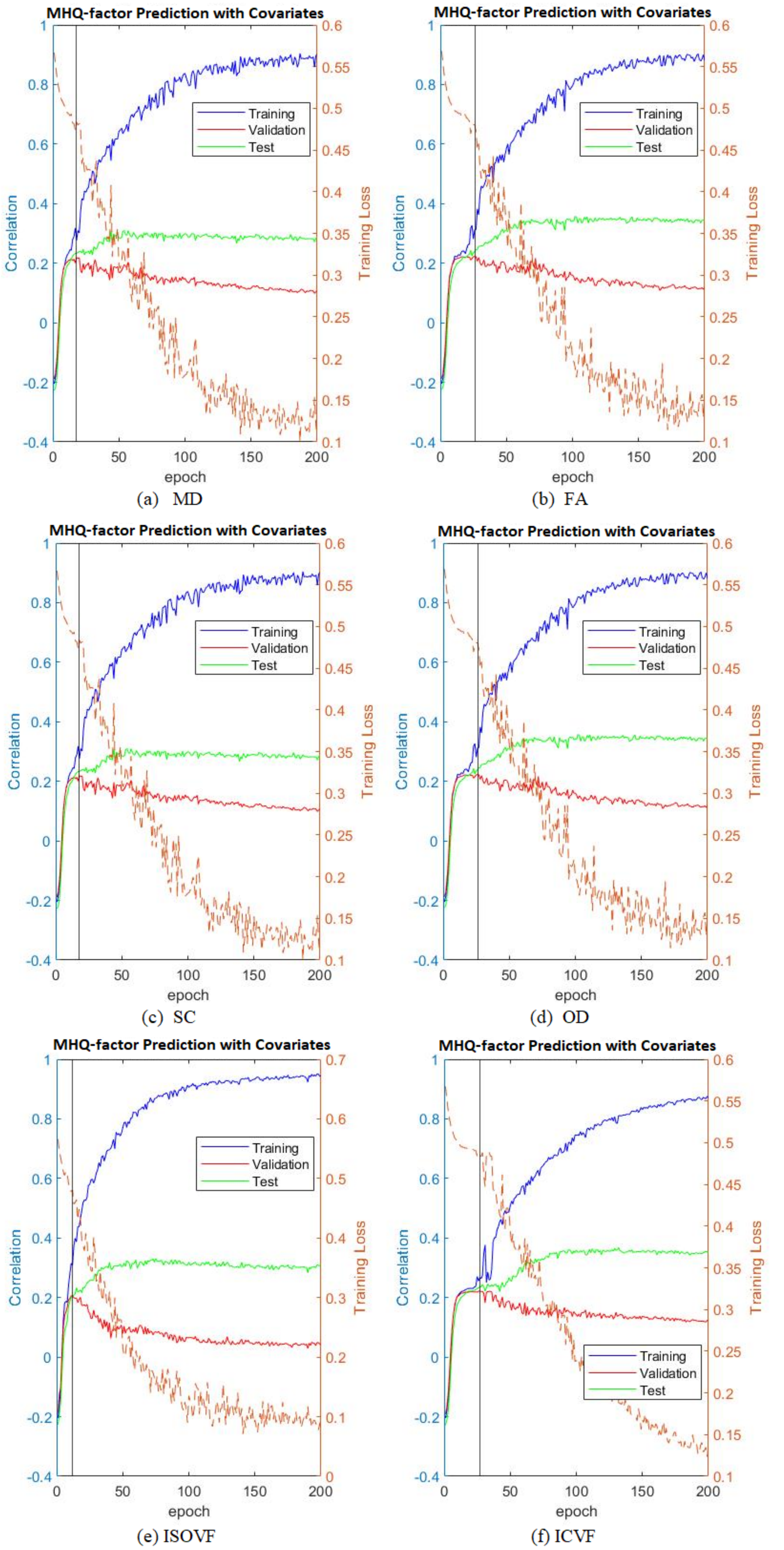


Figure 9: The BrainNetCNN models' training progress for MHQ-factor prediction for the fourth fold based on the six different dMRI measures. Training loss is the cross-entropy loss
